# ClustALL: A robust clustering strategy for stratification of patients with acutely decompensated cirrhosis

**DOI:** 10.1101/2023.11.17.23298672

**Authors:** Sara Palomino-Echeverria, Estefania Huergou, Asier Ortega-Legarre, Eva M. Uson, Ferran Aguilar, Carlos de la Pena, Cristina Lopez-Vicario, Carlo Alessandria, Wim Laleman, Alberto Farias Queiroz, Richard Moreau, Javier Fernandez, Vicente Arroyo, Paolo Caraceni, Vincenzo Lagani, Cristina Sanchez, Joan Claria, Jesper Tegner, Jonel Trebicka, Narsis Kiani, Nuria Planell, Pierre-Emmanuel Rautou, David Gomez-Cabrero

**Author notes:** Contributed equally (Nuria Planell, Pierre-Emmanuel Rautou, David Gomez-Cabrero).

## Abstract

Patient heterogeneity represents a significant challenge for both individual patient management and clinical trial design, especially in the context of complex diseases. Most existing clinical classifications are based on scores built to predict patients’ outcomes. These classical methods may thus miss features that contribute to heterogeneity without necessarily translating into prognostic implications.

To address patient heterogeneity at hospital admission, we developed ClustALL, a computational pipeline designed to handle common clinical data challenges such as mixed data types, missing values, and collinearity. ClustALL also facilitates the unsupervised identification of multiple and robust stratifications. We applied ClustALL to a prospective European multicentre cohort of patients with acutely decompensated cirrhosis (AD) (n=766), a highly heterogeneous disease. ClustALL identified five robust stratifications for patients with AD, using only data at hospital admission. All stratifications included markers of impaired liver function and number of organ dysfunction or failure, and most included precipitating events. When focusing on one of these stratifications, patients were categorized into three clusters characterized by typical clinical features but also having a prognostic value. Re-assessment of patient stratification during follow-up delineated patients’ outcomes, with further improvement of the prognostic value of the stratification. We validated these findings in an independent prospective multicentre cohort of patients from Latin America (n=580).

In conclusion, this study developed ClustALL, a novel and robust stratification method capable of addressing challenges tied to intricate clinical data and applicable to complex diseases. By applying ClustALL to patients with AD, we identified three patient clusters, offering insights that could guide future clinical trial design.

## Introduction

Heterogeneity is a prevalent phenomenon observed in numerous diseases, including various types of cancer (1), autoimmune conditions like multiple sclerosis (2), and diabetes (3)). This becomes especially critical in diseases where environmental and lifestyle factors play a significant role. Acutely decompensated cirrhosis, which refers to the rapid development of overt ascites, overt hepatic encephalopathy, variceal haemorrhage, or any combination of these disorders, which often leads to nonelective admission to the hospital of patients who were previously stable (4), exemplifies significant inter-individual variability. It encompasses a range of causes of cirrhosis, comorbidities, precipitating events, clinical presentations, and outcomes (4). This clinical heterogeneity poses a considerable challenge as it likely accounts for the diverse responses to treatment and outcomes observed in these patients (5). Therefore, we reasoned that analysing a large population of patients with acutely decompensated cirrhosis should allow us to develop stratification tools.

A major tool for the characterization of patient heterogeneity is the identification of patient subtypes, also defined as patient stratification. Importantly, the World Health Organization has acknowledged patient stratification as a valuable approach for enhancing population health management and providing better-tailored services (6). In conceptual terms, patient stratification can be described as the process of grouping or clustering patients based on specific characteristics or patterns without relying on labelled data or information about future outcomes (7). Therefore, contrary to scores developed using classical statistical approaches based on the clinical course, stratification can capture features explaining patients’ heterogeneity independently of their association with patient outcomes.

Numerous attempts have been made to identify subgroups within clinical datasets (7–9). However, the lack of a universally applicable approach poses a significant challenge in the field of clustering analysis. Although there have been advancements beyond the classical k-means and hierarchical clustering methods, no general framework still allows the organization and classification of clustering methodologies in the clinical setting (10). Instead, many ad-hoc applications have been developed for specific scenarios, but their generalizability is often limited. While there is no global classification, these applications can be grouped based on specific characteristics such as managing missing values, collinearity, or mixed data (9). For instance, when handling missing data, some methods exclude samples from the analysis, potentially resulting in a loss of statistical power, while others rely on a single imputation, overlooking the potential bias that can be introduced (11). Highly correlated variables represent a challenge. Some methods exclude them, while others employ dimensionality reduction techniques such as Principal Component (PC) reduction to capture underlying lower-dimensional data patterns (12,13). However, both decisions may affect the outcome of the clustering, as sensitivity analyses are rarely conducted. Moreover, indiscriminate feature selection can inadvertently remove informative features along with noisy ones, potentially biasing the results (14). Furthermore, most clustering methodologies assume the existence of a single stratification, disregarding the possibility of having none or multiple valid alternatives for subgrouping the population (15). Interestingly, trace-based clustering methodologies have recently emerged to aid in the interpretation and validation of the identified subgroups, often requiring domain knowledge and expert input (16).

Additionally, the evaluation of clustering outcomes is an open problem that is based on the quality of the produced clusters. In the case of unsupervised clustering, where no preliminary classification exists, evaluations are typically referenced against theoretical benchmarks. For instance, when addressing the optimal number of clusters, various theoretical quality metrics are available such as the clustering coefficient (17) or the silhouette index (18) among many others. Importantly, while there is no universal methodology that excels across all scenarios for all data sets, as dictated by the “no free lunch” theorem (19), there exist strategies that yield high-quality results (20–22). Another essential measure—referred to as robustness—lacks a precise definition. Robustness, in general terms, signifies the capacity of a system to withstand changes (23). In our context, we investigate whether a clustering remains stable when subjected to perturbations. In this work, we considered two types of perturbations: those derived from changes in the population and those arising from changes in the algorithm’s parameters. In the case of population-based perturbations, we quantify how a given clustering is influenced by variations in the underlying population. Bootstrapping is one approach to address this scenario (24). In the case of parameter-based perturbations, we assess the impact of parameter adjustments in the clustering algorithm on the identified clustering (25). Consider a scenario where a parameter “x” defines our clustering strategy. How different is the resulting clustering when using “x=1” versus “x=1.1”? Here, robustness translates to clusterings that maintain stability even when parameter values shift. For the reader’s clarity, we will name the two different robustness criteria: population-based robustness and parameter-based robustness.

Importantly, there is currently no methodology capable of addressing all the aforementioned scenarios while ensuring both definitions of robustness. To address these challenges comprehensively, we developed ClustALL, a novel framework that robustly identifies patient subgroups by addressing all the previously mentioned challenges and limitations of existing methodologies, and applied ClustALL-as a proof-of-concept-in two large prospective cohorts of patients non-electively admitted to the hospital for acutely decompensated cirrhosis.

In this study, ClustALL was applied to a large prospective cohort of patients non-electively admitted to the hospital for acutely decompensated cirrhosis. The resulting stratifications were thoroughly characterized, aiming to identify any particular stratification of special interest in the clinical context showing prognostic value. We then validated the reproducibility of this stratification using a separate prospective cohort of patients. One further aim of the study was to demonstrate the usability of stratification over the disease course, with prognostic value.

## Results

### ClustALL, a robust data-driven framework for patient stratification in complex diseases

We developed a specialized stratification framework, referred to as ClustALL, specifically designed to accurately identify all potential alternatives for stratifying a population using clinical multimodal data at hospital admission as input. The ClustALL methodology consists of three main steps illustrated in Figure 1 and detailed in the Methods section: (1) Data Complexity Reduction (depicted in the Green Panel of Fig.1) aims to simplify the original dataset by mitigating the impact of redundant information (highly correlated variables). As a result, we obtain a set of embeddings, each one derived from different groupings of clinical variables. (2) Stratification Process (depicted in the Purple Panel of Fig.1), where, for each embedding, multiple stratification analyses are performed using different combinations of distance metrics and clustering methodologies. From each combination, denoted as “embedding + distance metric + clustering method”, a stratification is derived. (3) Consensus-based Stratifications step (depicted in the Red Panel of Fig.1) aims to identify robust stratifications that, in addition, exhibit minimal variation when combination parameters (“embedding + distance metric + clustering method”) are slightly modified. ClustALL performs a population-based robustness analysis for each stratification using bootstrapping. This analysis ensures that combinations associated with non-robust stratifications are excluded. The resulting stratifications are then compared using the Jaccard distance. As a result, a heatmap is generated to visually identify groups of representative stratifications (green squared lines). The selection of representative stratifications enables the preservation of those stratifications that demonstrate parameter-based robustness: consistency even when various parameters, like distance metrics or clustering methods, are altered. For each group of stratifications, the centroid is selected as the final stratification (green squares).

**Figure 1.**
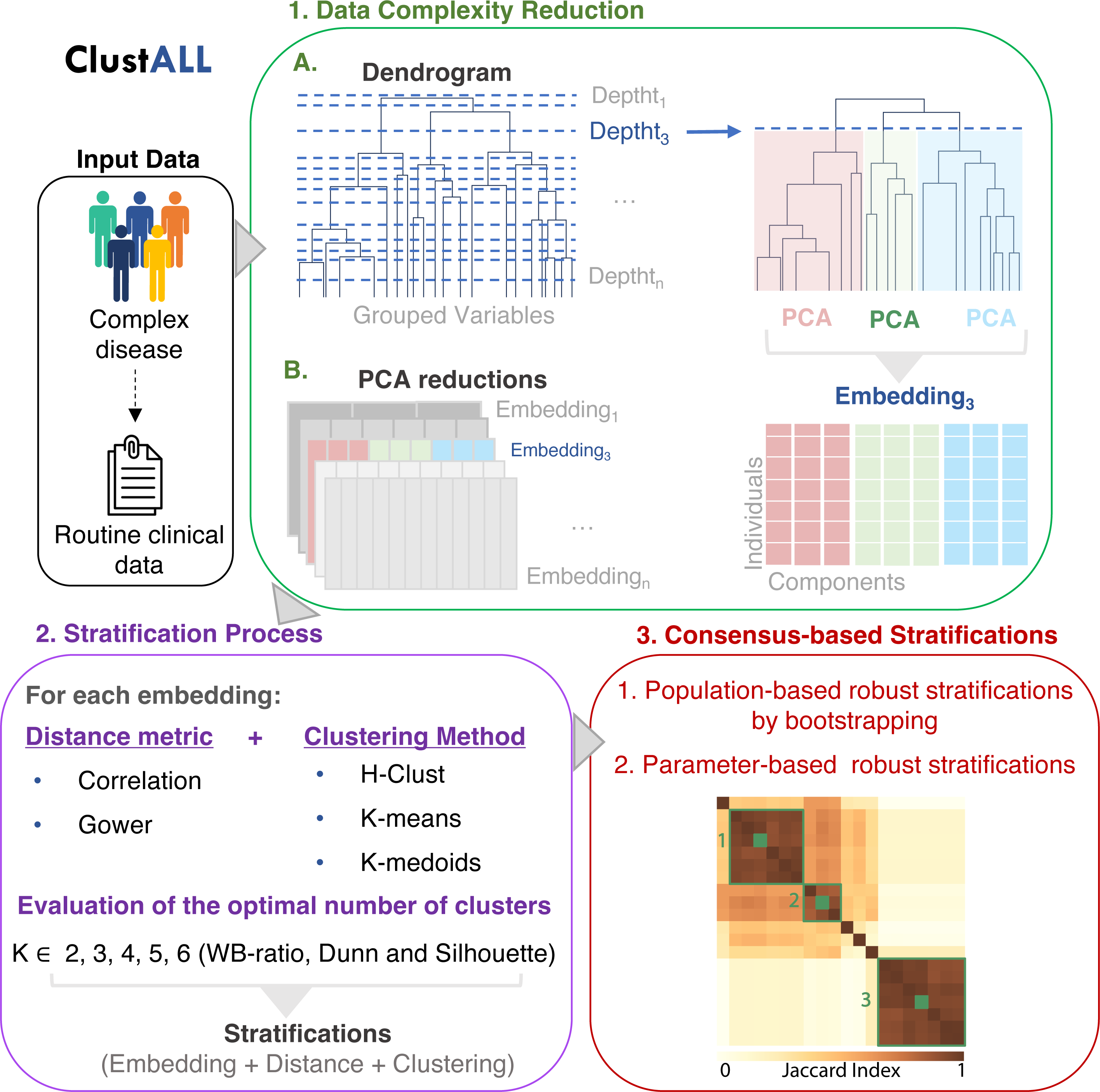
Schematic overview of the different steps of ClustALL approach (best viewed in colour). ClustALL takes clinical variables as input. First, data complexity is reduced by grouping the features into a dendrogram, assessing the resulting depths, and using Principal Component Analysis (PCA) (green panel). The output is an embedding for each possible depth. Then, stratification is computed considering the combination of different distance measures, clustering techniques, and cluster numbers (K) (purple panel). In the final step, non-robust stratifications are filtered, and the centroids derived from computing Jaccard (coloured green squares) similarity among the robust stratifications (green squares) are considered the final representatives of the stratifications (red panel).

Combining these three steps allows ClustALL to identify none, one, or multiple robust stratifications in a given population of patients with complex diseases. Importantly, a specific implementation of ClustALL is designed to effectively handle datasets with missing data effectively, ensuring that incomplete information does not hinder the stratification process.

### ClustALL uncovers stratification in a cohort of patients with acutely decompensated cirrhosis: a proof-of-concept

#### Study population

The ClustALL approach was applied to a subset of individuals from the European PREDICT cohort (26), which included 766 patients with acute decompensation of cirrhosis and 74 clinical features collected at hospital admission, with less than 30% missing values. Complete information on patient characteristics and short-term outcomes, including acute-on-chronic liver failure (ACLF), liver transplant, and death, can be found in Supplemental Table 1.

### ClustALL identified five different alternatives to stratify the population

The ClustALL workflow was utilized to discover potential new sub-phenotypes of patients with acute decompensation of cirrhosis within the PREDICT cohort upon hospital admission (Fig.2). To handle missing values in the dataset, we employed the ClustALL framework, which incorporates imputations using 1,000 iterations, as described in the Methods section. The Data Complexity Reduction Step resulted in 72 embeddings (Fig.2.1). The Stratification Process generated 288 stratifications based on the different combinations of “embedding + distance metric + clustering method” (Fig.2.2). Among these, 144 population-based robust stratifications were identified through the Consensus-based Stratifications step, resulting in five groups of parameter-based representative stratifications. The centroid was selected for each group of stratifications, (Fig.2.3).

**Figure 2.**
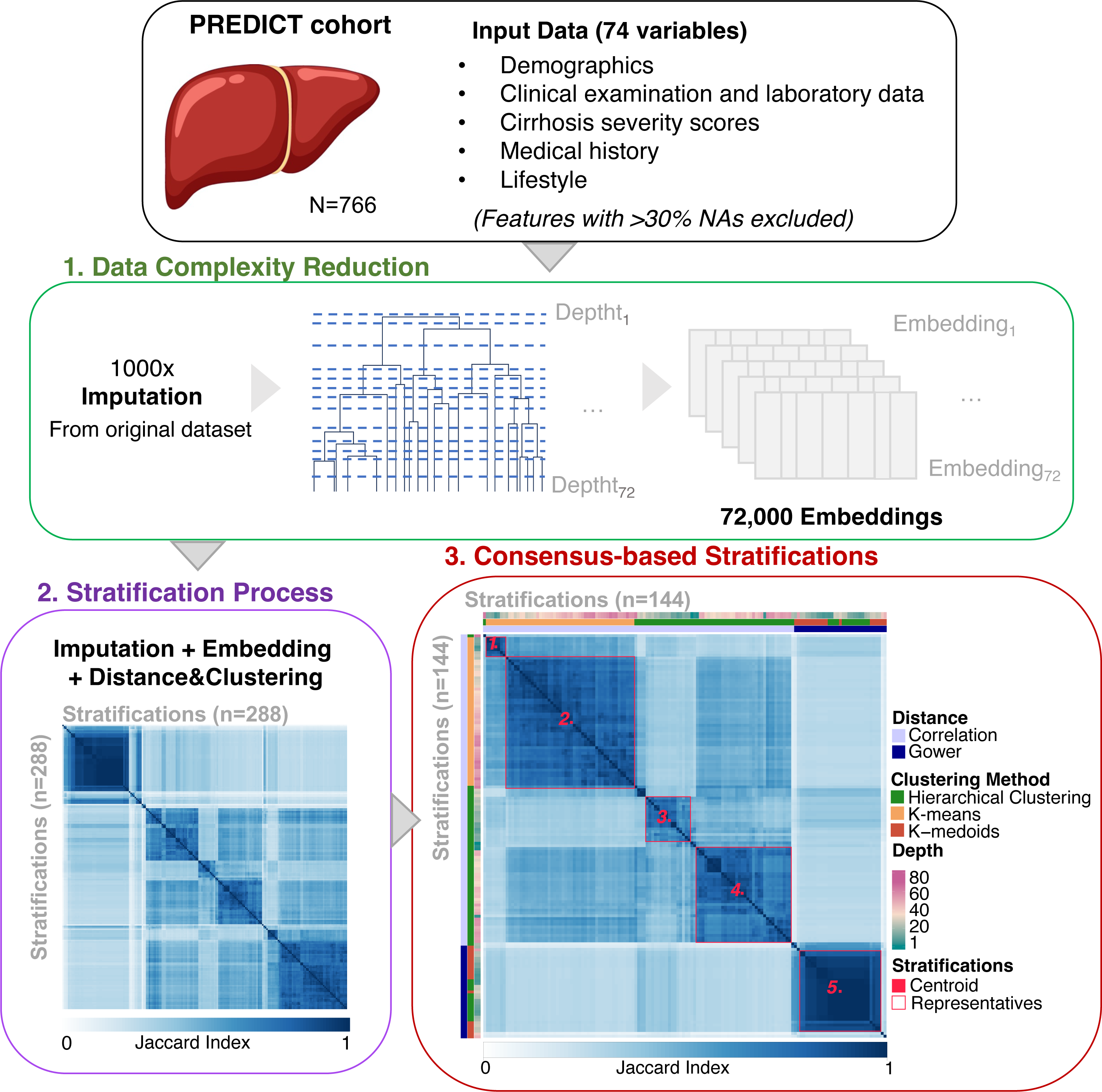
Summary of the outputs from the different steps of the ClustALL framework when applied to the PREDICT cohort (N=766). Input data comprised 74 clinical features with less than 30% missing values. The analysis utilized 1,000 imputed datasets. The Data Complexity Reduction step (green) was applied to 72 depths of the 1,000 imputed datasets. The Stratification Process step (purple) considered various clustering combinations resulting in 288 stratifications. After bootstrapping, 144 robust stratifications remained. Finally, in the Consensus-based Stratification step (red), five groups of robust stratifications (red squares) were identified, and the centroid was selected from each group as the final stratifications (red coloured squares).

### ClustALL provides better resolution than classical clustering tools

We conducted an analysis to assess the added value of ClustALL when compared with classical clustering methodologies such as k-means or hierarchical clustering. Regarding the classical methodologies, our findings revealed that when using correlation as a distance metric, 90% of patients were consistently assigned to a single cluster, regardless of the number of clusters considered; when Gower distance was utilized, the distribution of patients across clusters presented a more balanced distribution (Table S2). Notably, the population-based robustness of the stratifications generated by ClustALL was significantly higher (p-value <0.01) compared to the results obtained using k-means and hierarchical clustering (Fig.S3). In summary, our observations demonstrate that ClustALL significantly outperforms classical methodologies regarding population-based robustness.

### Characterization of the five robust stratifications within the PREDICT population

After identifying the robust stratifications, we aimed to explore and characterize the distinct clusters observed in each of the five alternative stratifications. These stratifications divided the patients into two clusters, except for stratification 1, which had three clusters. We visually investigated the separation by representing each stratification in a low-dimensional space using the corresponding embeddings derived from the dendrogram depths (Fig.3A-E) and the complete dataset (Fig.S2). Further exploration revealed that stratification 1 was a subdivision of stratification 2 (Fig.3F). We then determined the minimal sets of variables (excluding the cirrhosis severity scores (Table S1 variables 44 to 48)) with the highest predictive performance in differentiating the clusters for each stratification (Tables S3-S7) (27). The different classification approaches were described by 25 variables from a total of 74 (Table S1 variables 1 to 74), with 8 to 12 variables per stratification (Fig.4A). Notably, all stratifications included: (i) serum bilirubin concentration (either as a continuous variable or categorized under the term “liver dysfunction”(28)) (ii) International Normalized Ratio (INR) (either as a continuous variable or categorized under the term “coagulation dysfunction” (28); (iii) the number of organ dysfunction or failure. Precipitating events were present in all but one stratification (stratification 3) either as a sum or individually (gastrointestinal bleeding, alcohol-related hepatitis, acute viral hepatitis). Diabetes mellitus was included in two stratifications. Conversely, age, sex, BMI, cause of cirrhosis and lifestyle were present in no or one stratification. Interestingly, stratification 1 and 2 shared almost the same minimal set of variables. Both stratifications identified a group of patients with a severe phenotype attested by low serum sodium, low serum albumin, high serum bilirubin, high INR, high C-Reactive Protein (CRP) and leucocytes, and the number of precipitating events (Fig.4B). Hepatic encephalopathy was present in stratification 1 but not in 2 (29). A complete statistical characterization of the stratifications is provided in Tables S3 to S7. Considering the clinical implications of the features and the finer classification of the patients, we identified stratification 1 as the most insightful for further exploration in patients with acute decompensation of cirrhosis. Henceforth, in our discussions, we will refer to this specific stratification as ‘AD-strat’.

**Figure 3.**
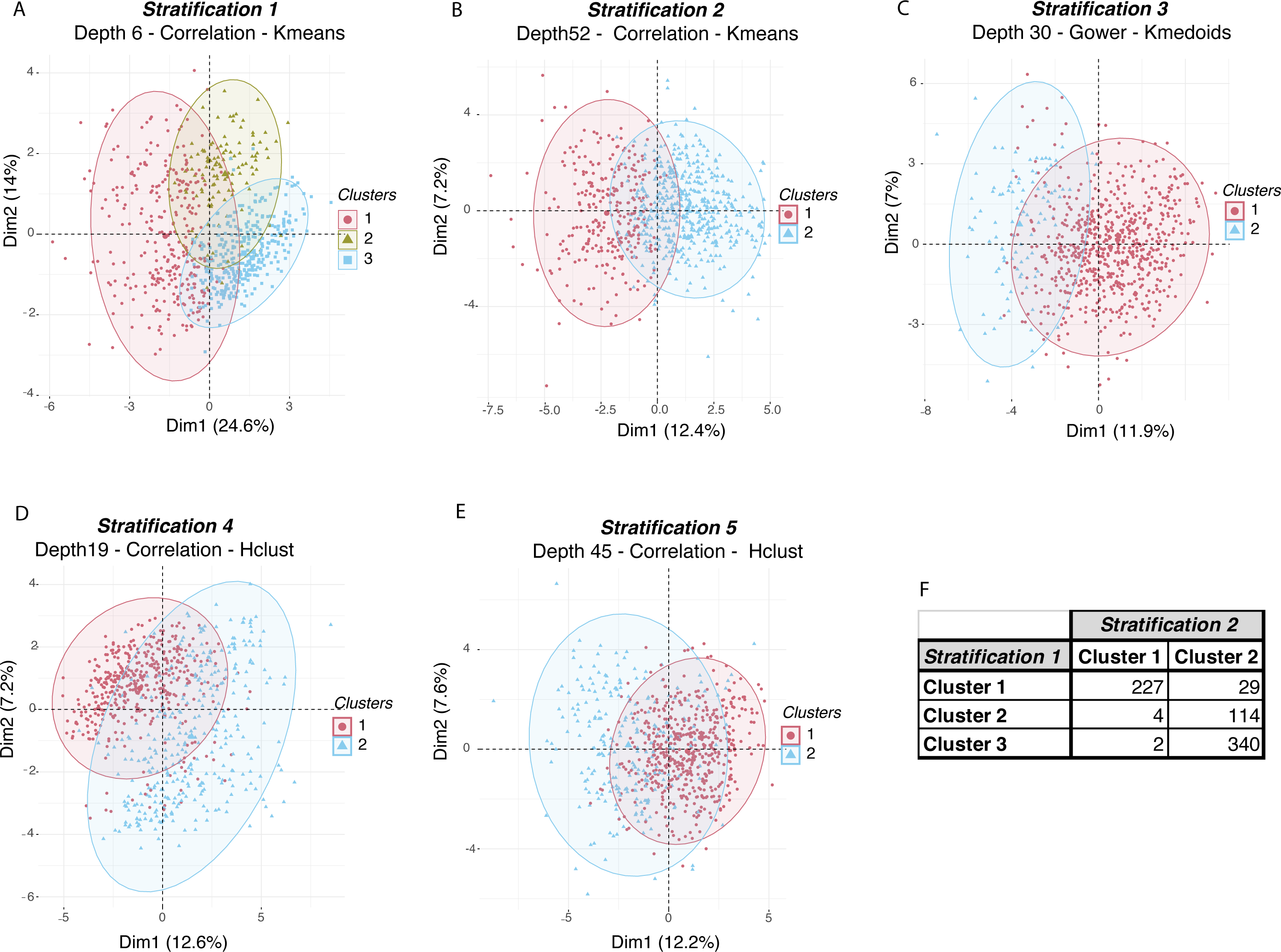
Principal Component projection of the ClustALL robust stratifications based on the embedding associated with each stratification. (A-E). Low-dimension representation of the robust stratifications after applying the ClustALL framework to the PREDICT cohort. For each one of the 5 robust stratifications identified by ClustALL, the Principal Component Analysis of the Embeddings corresponding to the specific dendrogram depth associated with the stratification is shown. The x (Dim1) and y (Dim2) axes represent the first and second principal components respectively, which are linear combinations of the original variables. (F). The overlap between the clusters in stratifications 1 and 2 shows that stratification 1 is a subdivision of stratification 2.

**Figure 4.**
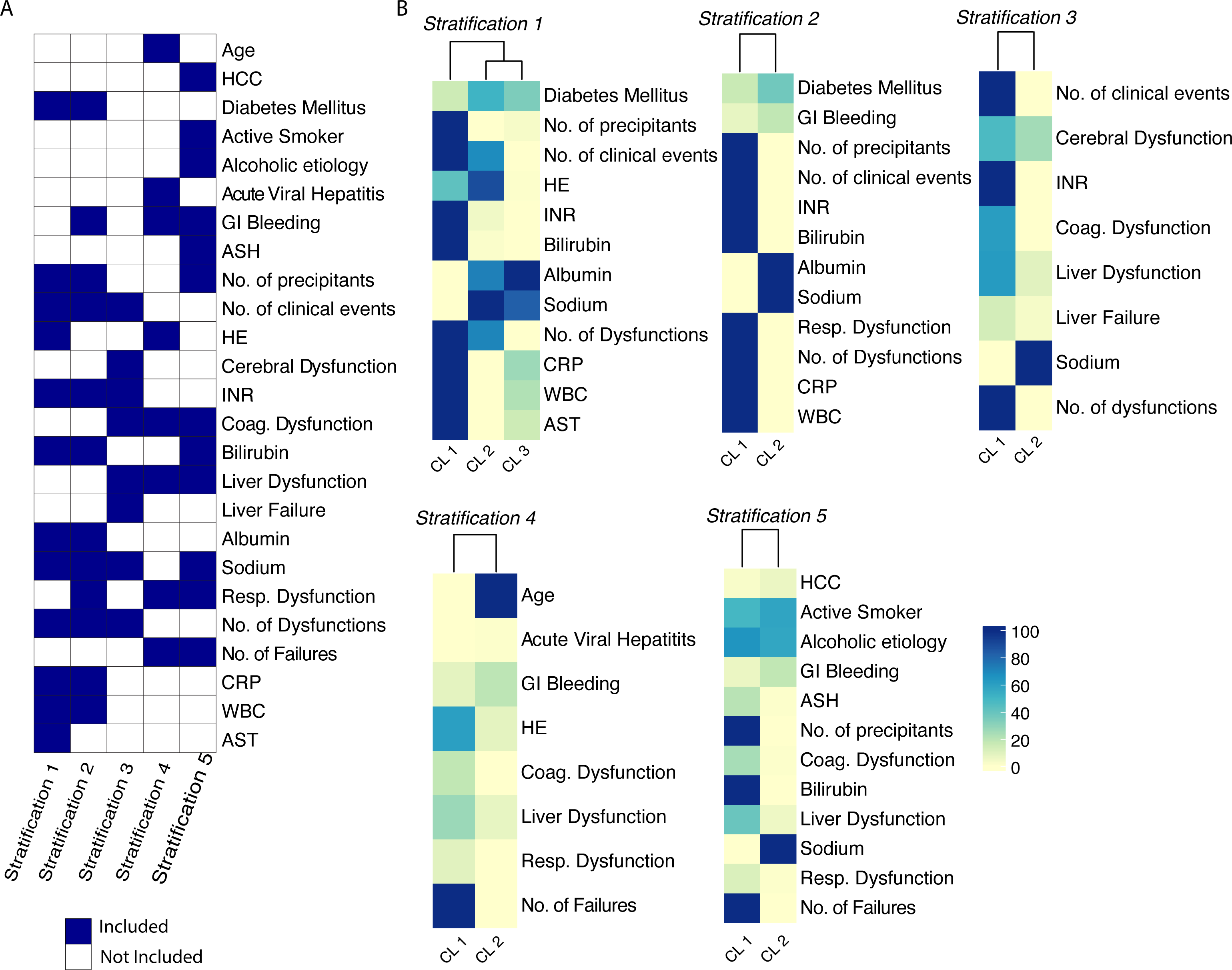
Overview of the variables driving the ClustALL stratifications. (A). Heatmap with the minimal set of variables required to describe the 5 different stratifications, accounting for 25 out of 74 input variables. (B). Heatmaps of the minimal set of patient characteristics per stratification. The heatmap colour scale depends on the data type. In the case of binary variables, the value indicates the percentage of patients with such binary characteristics, e.g., the presence of Diabetes Mellitus. For continuous variables, the colour scale represents a scaled value from the highest cluster mean (100.0) to the lowest cluster mean (0.0), e.g., Albumin and CRP. Abbreviations: ASH = Acute Alcoholic-Steatohepatitis, AST = Aspartate aminotransferase, CL= Cluster, CRP = C-Reactive Protein, HE = Hepatic encephalopathy, HCC = Hepatocellular Carcinoma, INR = International normalized ratio, WBC = White blood cell counts.

### *AD-strat* provides prognosis value

The AD-strat stratification is defined by three subgroups (clusters) of patients with acutely decompensated cirrhosis revealing different clinical characteristics and disease progression. Cluster 1 included 306 patients (39.95%) who exhibited the most clinically critical scenario (Fig.5A, B and Table S3). These individuals had the highest rates of organ dysfunction, clinical events, and precipitating events (Table S1). They had a marked acute inflammatory profile (high white blood cell count, and C-reactive protein level) poor liver function (low levels of albumin, and high levels of INR and serum bilirubin) and more hepatocyte injury (higher levels of serum aspartate aminotransferase). Conversely, Cluster 2 (n=118; 15.4%) and Cluster 3 (n=342; 44.6%) had a less severe presentation. The main difference between Cluster 2 and 3 was hepatic encephalopathy, found in 89% of the patients in Cluster 2 and almost no patients in Cluster 3 (Fig.5A, B and Table S2). Importantly, a significant prognostic value of AD-strat was revealed by exploring the cumulative incidence of ACLF and death over 90-day follow-up (Fig.5C). Patients in Cluster 1 had poor short-term outcome, with a cumulative incidence of ACLF and death, both by 90 days of 24.1 and 21.5, respectively. While Clusters 2 and 3 had similar risks of ACLF by 90 days (8.6% and 10.2 %, respectively), the risk of death by 90 days was lower for Cluster 2 than Cluster 3 (4.3% vs 10.7%).

**Figure 5.**
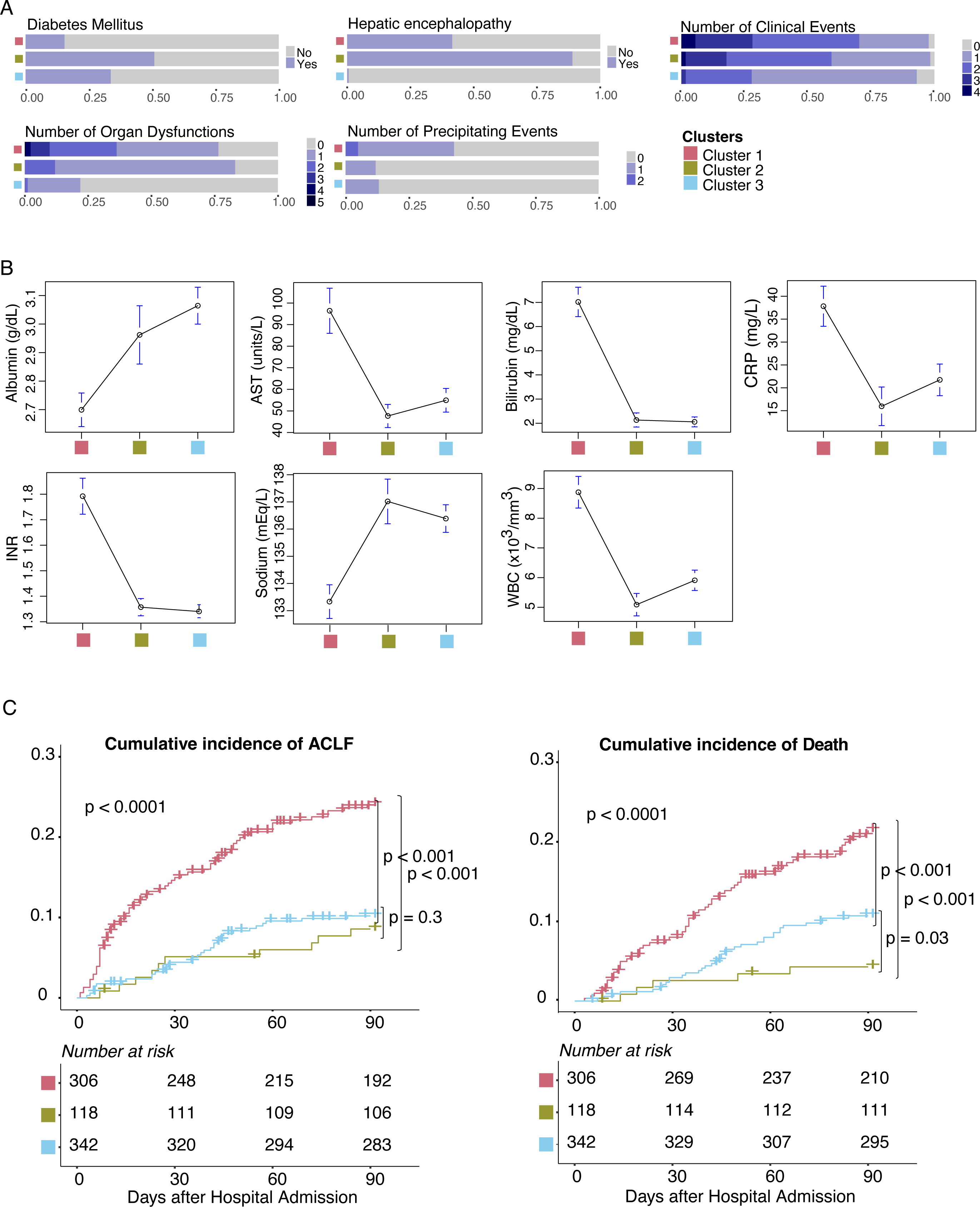
Clinical overview of the AD-strat derived clusters in the PREDICT cohort. (A, B). Distribution of the highest predictive performance-related patient characteristics among AD-strat clusters; (A) categorical variables, (B) numerical variables. C) Cumulative incidence of ACLF (left) and death (right) according to the AD-strat clustering in PREDICT cohort considering 90 days after hospital admission, with the number of patients at risk per cluster (Transplantation counted as a competing risk to death). Abbreviations: AST = Aspartate aminotransferase, CRP = C-Reactive Protein, INR = International normalized ratio, WBC = White blood cell counts.

When we compared the clusters identified with ClustALL - exclusively using data obtained at admission - with the groups of patients based on their clinical course (26), we found a statistically significant association (Fisher test, p-value < 0.01) (see Table S8). We observed that 61% of patients with pre-ACLF were in Cluster 1 and 48% of patients with stable decompensated were in Cluster 3.

### Reproducibility of the stratification model in an independent cohort

We assessed the validity of the AD-strat model in a large independent prospective multicentre cohort that included 580 patients with acute decompensation of cirrhosis from the Latin-American ACLARA study (30). Using as a reference the PREDICT AD-strat clusters, we labelled ACLARA patients using the k-nearest neighbours (kNN) classification algorithm (Table S9) (31). The classification model included the 12 predictive variables previously identified in the feature importance analysis (Fig.3B Stratification 1). Importantly, the labelling was consistent and independent of the imputation (Fig.6A), and the distribution of individuals by AD-strat clusters within ACLARA closely mirrored that of the PREDICT cohort (Fig.6B). As expected, the clustering of the ACLARA cohort exhibited similar clinical feature patterns to the PREDICT cohort (Fig.6C, Fig.3B Stratification 1). Furthermore, the features describing the subgroups demonstrated statistical significance (Table S10). Finally, we assessed the clinical relevance of the clustering in terms of prognosis, specifically examining the short-term outcomes available in the ACLARA cohort 28 days after hospital admission. Similar to results obtained in the PREDICT cohort, Cluster 1 displayed a bad prognosis for both ACLF and death, while Cluster 3 showed a better prognosis (Fig.6D). In ACLARA, all patients from Cluster 2 were afflicted by hepatic encephalopathy (Table S10) and showed a poor prognosis similar to that of Cluster 1. Ethnicity was homogeneously distributed across clusters (Table S2). In particular, Native Americans represented 21% of Cluster 1, 15% of Cluster 2, and 14% of Cluster 3. Complete information on patient characteristics and short-term outcomes is reported in Supplemental Table 9.

**Figure 6.**
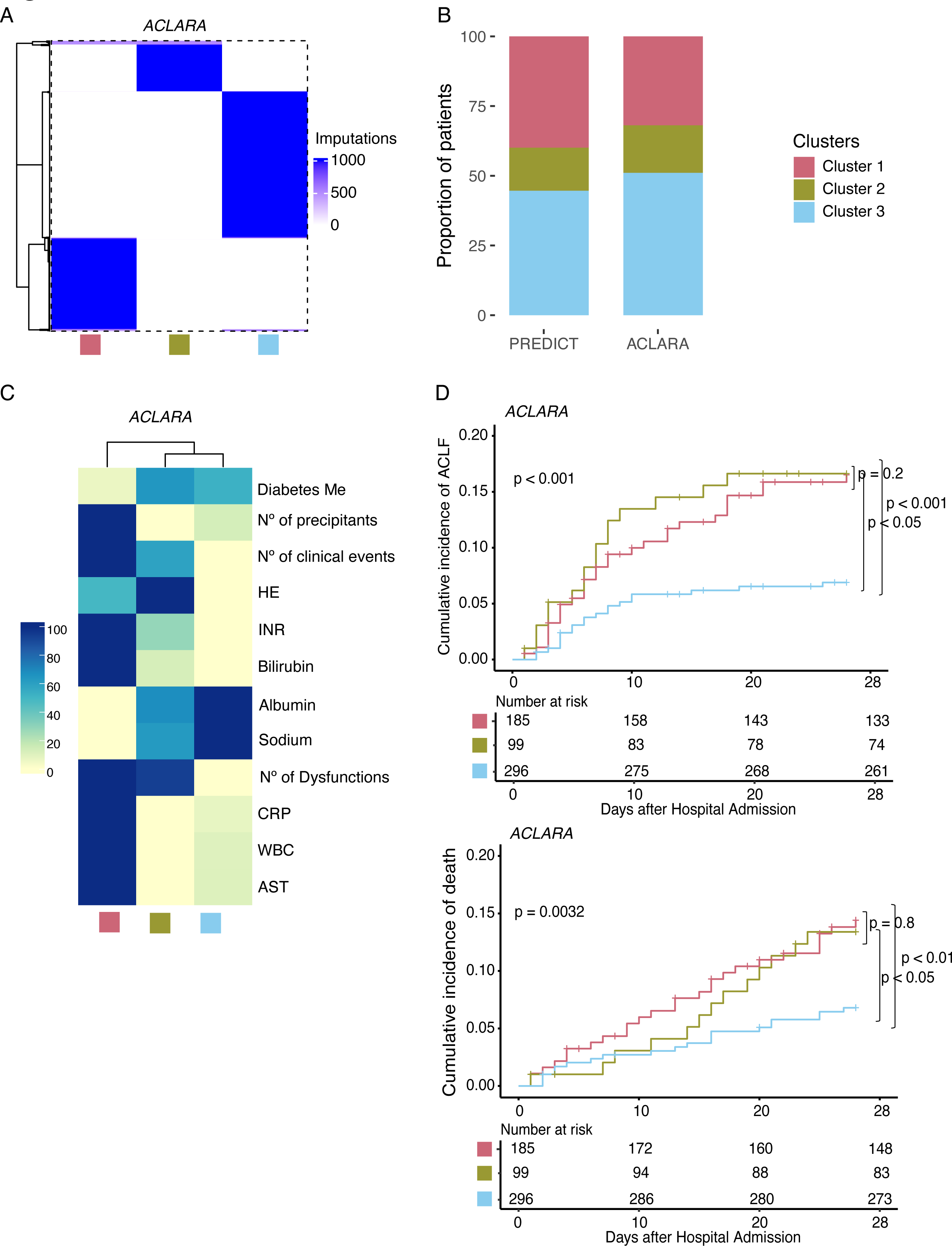
Reproducibility of the AD-strat model in the ACLARA cohort. (A) Distribution of the labels in the ACLARA cohort after applying the kNN model 1,000 times. (B) Proportion of patients distributed in the 3 clusters in the PREDICT and the ACLARA cohorts. (C) Heatmap of patient characteristics per cluster in the ACLARA cohort. Bars on the right show the colour scale representing the proportion with each binary characteristic, such as diabetes. Continuous variables, such as bilirubin, represent a scaled value from the highest cluster mean (1.0) to the lowest cluster mean (0.0). (D) Cumulative incidence of ACLF (up) and death (down) according to the AD-strat clustering in ACLARA cohort considering 28 days after hospital admission, with the number of patients at risk per cluster (Transplantation counted as a competing risk to death). Abbreviations: AST = Aspartate aminotransferase, CRP = C-Reactive Protein, INR = International normalized ratio, WBC = White blood cell counts.

### *AD-strat* as a marker for clinical management

Finally, we investigated the clinical value of the stratification during the follow-up visits of the PREDICT cohort. Based on the PREDICT study design (26), two follow-up visit plans were established depending on the reported disease severity (CLIF-C AD-score) at hospital admission (Fig. 7A). For patients with a CLIF-C AD-score ≥ 50, the scheduled visits were performed at hospital admission and 1, 4, 8 and, 12 weeks after enrolment. For patients with a CLIF-C AD-score < 50, the scheduled visits were performed only at hospital admission and 1 and 12 weeks after enrolment.

**Figure 7.**
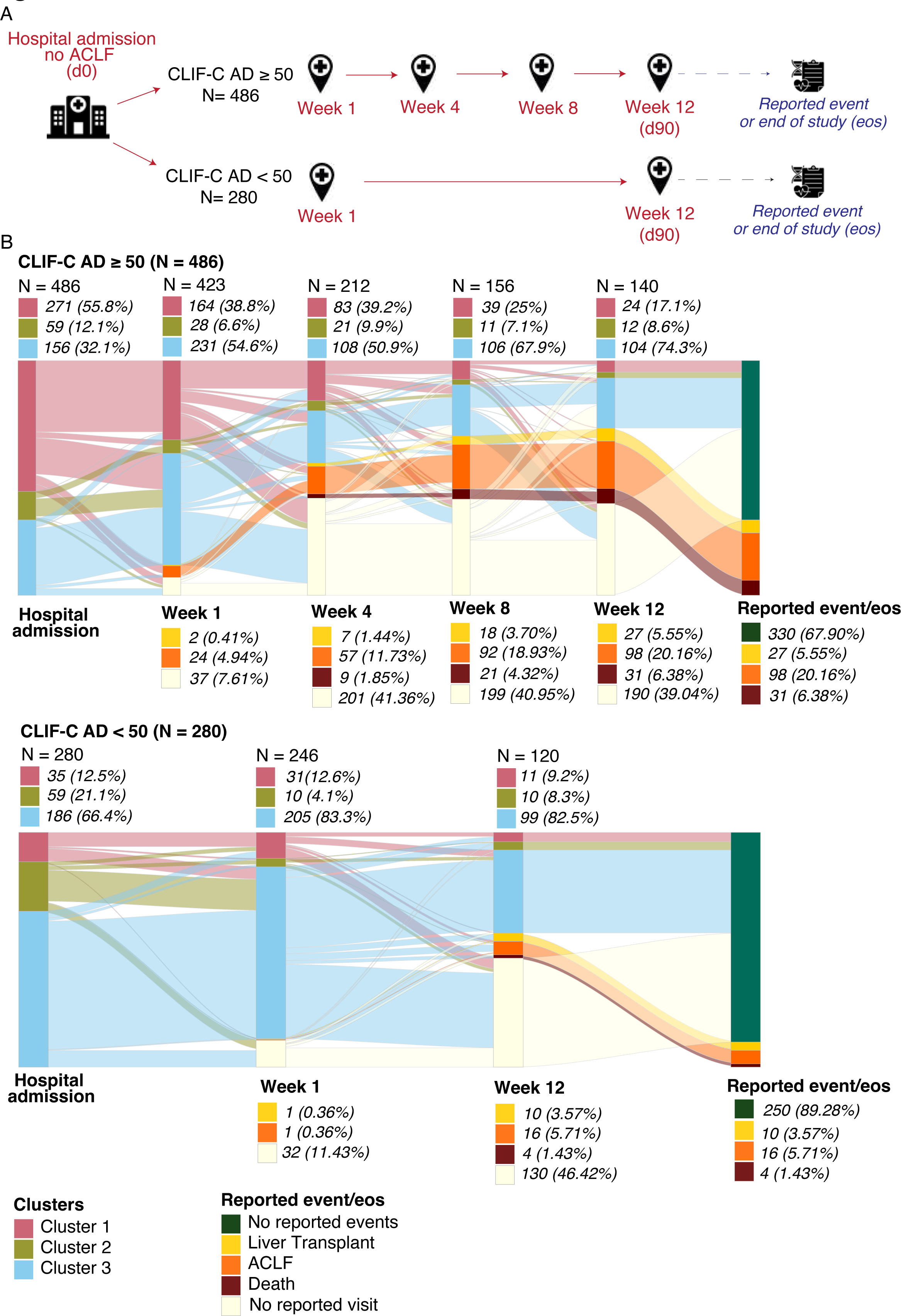
Distribution and transition of the AD-strat derived clusters at different visits in the PREDICT cohort. (A). Schematic representation of PREDICT study design. Two follow-up visit plans were defined according to the reported disease severity (CLIF-C AD-score) at hospital admission (red). The information about the occurrence of any adverse event (liver transplant, ACLF or death) during the whole visit plan or the absence of events at the end of the study was tracked (blue). (B) Sankey plots show the cluster label of each patient over the follow-up visits. The follow-up flows of patients with CLIF-C AD >= 50 at hospital admission (up) and CLIF-C AD <50 at hospital admission (down) are shown. The distribution of the patients assessed at each follow-up visit per cluster is shown as frequency and proportion on the top of the Sankey representations. The accumulated frequency and proportion of adverse events at each follow-up visit respecting the whole cohort (for CLIF-C AD >=50, n=486; for CLIF-C AD < 50, n=280) are shown on the bottom of the Sankey representations. Reported event/eos, shows the status of a patient at the “end of the study”: patients with a reported event or patients with no reported event.

Of the 766 patients included in the PREDICT study, 688 had at least one follow-up visit. For this subset of patients with available data, we labelled each of them at each follow-up visit using the kNN algorithm (Fig. 7B). This approach allowed an overview of the patient stratification over the entire study duration and revealed the patient flow over time highlighting cluster transitions.

Consistent with the previous AD-strat characterization at hospital admission (Fig.5 and Table S3), we identified more than 50% of patients with CLIF-C AD score ≥ 50 (n=486) were classified as Cluster 1, while patients with CLIF-C AD score < 50 (n=280) were predominantly classified as Cluster 3 (66.4%) (Fig.7B). Changes in cluster proportions were observed during the patients’ follow-up. Stratification changes over time were more pronounced among patients with CLIF-C AD scores≥ 50 at hospital admission, showing a progressive reduction of patients classified as Cluster 1 (55.8% at HA, 38.8% at week 1, 39.2% at week 4, 25% at week 8, and 17.1% at week 12) and an increase of those classified as Cluster 3 (32.1% at HA, 54.6% at week 1, 50.9% at week 4, 67.9% at week 8, and 74.3% at week 12). Additionally, there was a progressive increase in the proportion of patients classified as Cluster 3 for those patients with CLIF-C AD-score < 50 at hospital inclusion (66.4% at HA, 83.3% at week 1, and 82.5% at week 12).

To assess the effectiveness of the AD-strat throughout disease progression, we determined its prognostic value in two scenarios: 1) using the stratification at hospital admission, and 2) using the stratification at the last visit reported before the occurrence of any adverse event (we considered any visit between week 1 and 12) or at the end-of-study (EOS) (week 12 visit). A significant difference was observed (p<0.001, Wilcoxon test) when comparing the time window between the visit used in each scenario and the occurrence of adverse events (Fig. S5), indicating that in the second scenario, we evaluated patients during a visit much closer to the event.

Ultimately, the cumulative incidence of ACLF and death as stratified at the last visit demonstrated a more significant separation between clusters compared to patient stratification at hospital admission (Fig.8). There was an increase in the incidence for those patients classified as Cluster 1 (18.46% and 18.45% at baseline and 28.16% and 26.8% at the last visit for ACLF and death, respectively). Accordingly, the goodness-of-fit parameters indicated an improvement in risk prediction with the last visit stratification, suggesting an enhanced predictive power as the event approached (Table S11).

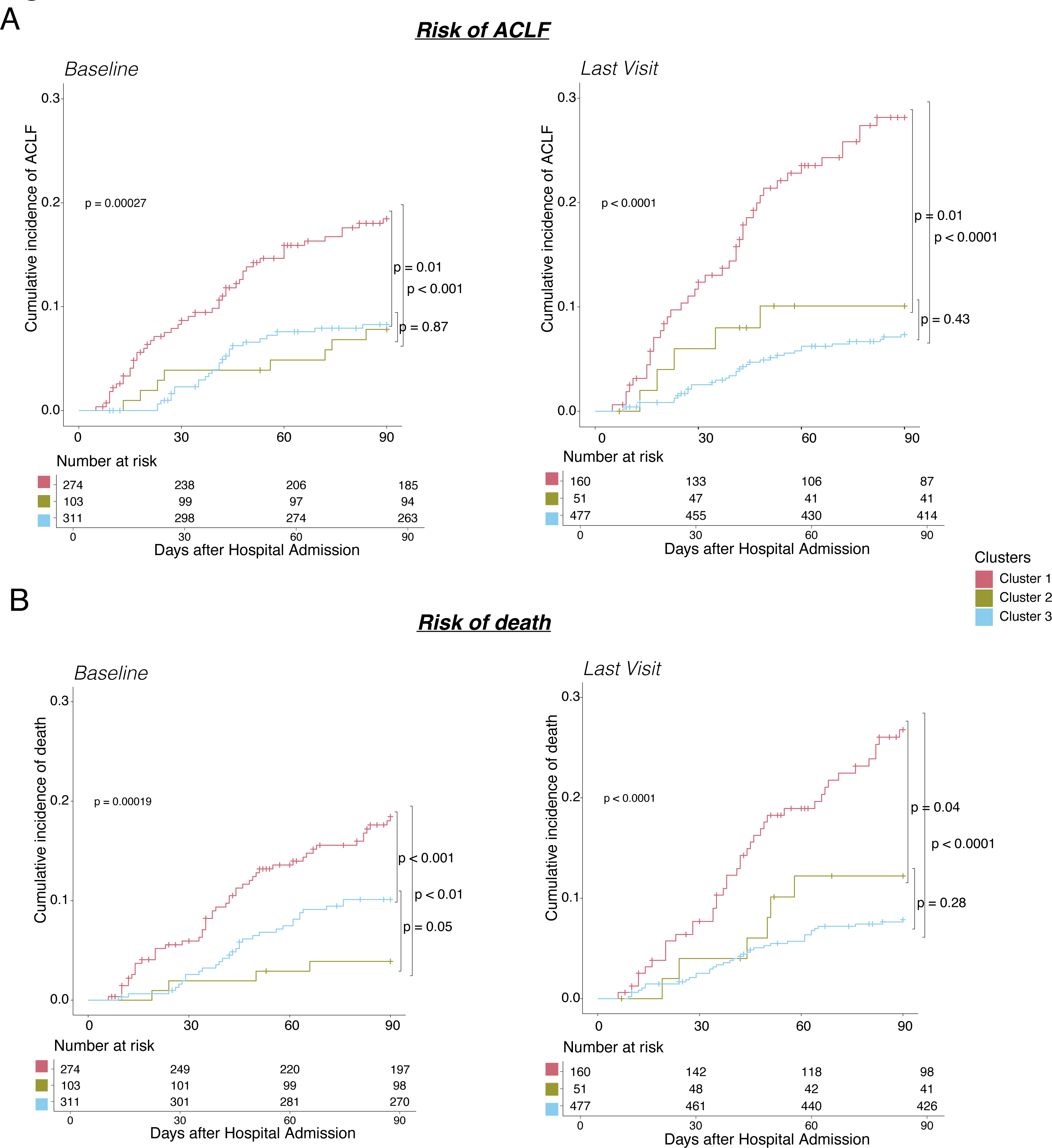

## Methods

### ClustALL Framework

Given a set of patients affected by a complex disease with clinical data available, the goal of ClustALL is to identify all the possible alternatives to stratify them that are robust and consistent, even when different parameters or settings are used to generate the stratifications (distance metric, clustering algorithm, and the number of imputations).

### Input data

ClustALL accepts both binary and numerical clinical variables as input. Categorical features are transformed using a one-hot encoder method. A minimum of two features is required, but including more features would lead to more precise clustering. It is important to note that increasing the number of features may also increase the computation time.

### Step 1. Data Complexity Reduction

In this step, highly correlated features are replaced by a reduced set of variables that account for their variability. To that end:

<u>Step 1.1. Dendrogram.</u> Hierarchical clustering is applied to the data, resulting in a dendrogram where variables are grouped based on similarity (32). The depth of each branch represents the distance between the groups of variables. All the possible depths of the dendrogram are extracted, and the sets of variables beneath each depth are stored as Depth.

<u>Step 1.2. Preprocessing</u>. Principal Component Analysis (PCA) is computed for each set of variables corresponding to each Depth, and the first three principal components are stored in a new matrix (Embedding) (33). For sets that contain only one variable, the variable itself is stored to generate the replacement matrix. This results in a complexity-reduced data set (Embedding) for each considered Depth. A subset of depths can be considered when the number of variables is too large.

### Step 2 Stratification Process

In this step, ClustALL calculates and pre-evaluates stratifications for each Embedding. For each Embedding, the dissimilarity between patients’ pairs is computed using correlation-based distance and Gower dissimilarity metric, resulting in two distance matrices. Clustering algorithms are then applied (34–36) depending on the distance used: k-means and hierarchical clustering for correlation distance matrices, and k-medoids and hierarchical clustering for the Gower distance matrix. Throughout all experiments, five different cluster numbers are evaluated kE{2, 3, 4, 5, 6}. The optimal number of clusters for each strategy is determined based on the consensus from three different measures of clustering internal validation: the sum-of-squares based index or WB-ratio, the Dunn index, and the average silhouette width (37,38). The objective is to group patients with comparable data while ensuring that patients in separate clusters are as dissimilar as possible from those in other clusters. As the output for this step, a stratification is derived for each combination denoted as “embedding + distance metric + clustering method”.

### Step 3 Consensus-based stratifications

<u>Step 3.1. Population-based robustness.</u> A data-driven threshold is used to define population-based robust subgroups or clusters. For each resulting stratification from the previous step, cluster-wise stability is computed by bootstrapping the dataset 1,000 times and calculating the Jaccard similarity index to the originally defined clusters (39). Stratifications with less than 85% stability (Fig.S4) are excluded based on data distribution. The remaining stratifications are denoted as Strat_filt_.

<u>Step 3.2.</u> Jaccard distance is applied to compute distances between the population-based robust stratifications. Then, to identify parameter-based robust clusters (where a minor modification in parameter selection provides a similar result), ClustALL considers those combinations that are part of a group of stratifications (green squares in Consensus-based stratifications step in Fig.1). Then, as initial criteria, that can be modified by the user, centroids from each “combination group” are selected as parameter-based robust stratifications (coloured green squares in Consensus-based stratifications step in Fig.1). The outcome can be none, one, or multiple ways to stratify the population robustly. In the current analysis, we considered parameter-based robust representatives: centroids of a combination group that includes at least 5 population-based robust stratifications.

### ClustALL enables input data with missing values

ClustALL can be adapted to work with missing data (Fig.S1). To that end, the ClustALL method is modified as follows:

<u>Step 1 adaptation</u>. First, a dendrogram and its associated depths are computed considering the original dataset with missing values. The original dataset is then imputed 1,000 times with the MICE algorithm to ensure the results are not derived from a single imputation (40). For each Depth previously calculated and each imputed dataset, the Data Complexity Reduction step is applied.

<u>Step 2 adaptation</u>. Step 2.1 is computed for each combination of depth, distance metric, clustering algorithm and each Embedding derived from an imputed dataset. The selection of the optimal number of clusters is based on the consensus from cluster internal validation and the mode of the imputed datasets for each corresponding embedding. Afterward, a distance matrix (D_mat_) between individuals is obtained by computing how often two individuals are assigned to the same cluster in each imputation (Fig.S1). Then, D_mat_ calculates a final stratification score using correlation-based distance and h-clust. In our experience, limited optimization is required here because summarizing the stratification over all imputations separately strengthens what is observed in each imputed dataset. Extra care will be required only in cases where imputations may differ significantly. After this modification, the method follows as previously described (Fig.S1).

### Data source

The data utilized in this study were obtained from two independent multicentre studies: the European PREDICT cohort and the Latin-American ACLARA cohort, conducted as part of the European Project DECISION (26,41). Both cohorts collected various measures including clinical, pharmacological, biomarker, and outcome data from patients with acute decompensation of cirrhosis upon hospital admission and during follow-up visits. The follow-up period was 90 days for the PREDICT cohort and 28 days for the ACLARA cohort. To be eligible for the present study, patients were required to have acute decompensation of cirrhosis upon hospital admission, with available information on short-term outcomes, drug intake, and available biological samples. Ultimately, 766 patients from the PREDICT cohort and 580 patients from the ACLARA cohort and 74 features (continuous and categorical) were included in the analysis. The features included demographic information, clinical and laboratory data, medical history, risk factors, and cirrhosis scores at hospital admission, with missing values accounting for less than 30% (Table S1). To avoid bias from missing data, imputation was performed with 1,000 iterations using the Multivariate Imputation by Chained Equations (MICE) method (21).

**Table 1.** Complete list of input features. Patient characteristics included in the analysis: demographics, cause of cirrhosis, main reason for hospitalization, manifestations at admission, cirrhosis severity scores, medical history, lifestyle and laboratory variables. Variables not included in ACLARA cohort.

|  |  |
| --- | --- |
| **Demographics** | Age, sex, height<sup>†</sup>, weight<sup>†</sup>, BMI, ethnicity (Black or African American, Asian, White, other) |
| **Cause of cirrhosis** | Alcohol, viral, alcohol + viral, NASH, cryptogenic, other |
| **Main reason for hospitalization** | Ascites, hepatic encephalopathy, gastrointestinal bleeding, spontaneous bacterial peritonitis, other infection |
| **Manifestations at admission** | Clinical Events (ascites, hepatic encephalopathy, gastrointestinal bleeding, acute kidney injury, bacterial infection, acute alcoholic-steatohepatitis, acute viral hepatitis, hepatocellular carcinoma), number of clinical events (the sum of clinical events), number of precipitating events (the sum of precipitating events: proven bacterial infection, acute alcoholic-steatohepatitis, CLIF-C AD > 50), organ dysfunction (liver, renal, cerebral, coagulation, cardiac, respiratory), number of organ dysfunctions (the sum of organ dysfunctions), organ failure (liver, cerebral, coagulation, cardiac, respiratory), number of organ failures (the sum of organ failures) |
| **Cirrhosis Severity Scores** | Child-Pugh, CLIF-C AD, CLIF-C OF, MELD, MELDNA |
| **Medical history** | History of diabetes, history of hypertension, history of previous decompensations |
| **Lifestyle** | Alcohol, active alcohol consumption<sup>†</sup>, tobacco |
| **Laboratory variables (measured in serum)** | Alanine transaminase, aspartate aminotransferase, albumin, bilirubin (total), gamma-GT, C-reactive protein, sodium, potassium, glucose<sup>†</sup>, hemoglobin, hematocrit, creatinine, white blood cell count, lymphocytes, monocytes, neutrophils, INR (International Normalized Ratio), platelet, SpO2 (%), SpO2/FiO2 Ratio |

### CLustALL comparison to different clustering methodologies

A comparison was conducted between the ClustALL framework and classical clustering algorithms. Stratification was performed on 1,000 imputed datasets using classical k-means and hierarchical methodologies with k values of 2 and 3, considering that ClustALL robust stratifications comprised two or three patient subgroups. Bootstrapping was performed for the classical clusters to evaluate cluster-wise stability (39). The resulting stability was compared to ClustALL stability through the Kolmogorov–Smirnov test. Moreover, the clinical utility of the various stratifications was assessed by examining the clinical insights obtained from the different clusters.

### Statistical Methods

All analyses were performed in the R Computing Environment version 4.0.3 (42).

### Descriptive statistics

Descriptive characteristics of the PREDICT and ACLARA study populations were reported as means with standard deviations for continuous variables and proportions of patients for categorical variables.

### Feature Analysis

The identification of the minimal-size predictive signatures with maximal predictive power leading to each stratification was performed using the fbed.reg function with default hyperparameters from the ‘MXM’ R package (27).

### Parametric Tests

Differences between clusters in the PREDICT and in the ACLARA cohorts were assessed using one-way ANOVA for continuous variables, while binary variables were tested with the chi-square test. The association between the PREDICT clusters identified with ClustALL - exclusively using data obtained at admission - with the groups of patients based on their clinical course (26), was tested with the Fisher test.

### Stratification model reproducibility

AD-strat model was validated in and in a separate cohort of patients with acute decompensation of cirrhosis from the ACLARA cohort and in PREDICT follow-up time points. For this purpose, the kNN model was trained on the PREDICT AD-strat cluster labels based on the signatures previously defined as most predictive in the feature analysis. The K parameter was selected based on accuracy, the area under the curve (AUC), error rate (ER), false positives (FP), and false negatives (FN) (Table S7). After applying the KNN algorithm, the target data (ACLARA cohort and PREDICT follow-up) was labelled based on the majority votes from the K nearest neighbours and imputed datasets.

### Survival Analysis

Cumulative incidences of ACLF development and liver-related death were estimated using the cumulative incidence function of the ‘survival’ R Package. Liver transplantation was considered a competing event. A p-value lower than 0.05 with Benjamini and Hochberg (BH) adjustment was considered statistically significant.

### Longitudinal analysis and model evaluation

All PREDICT patients with ≥1 post-baseline assessment (n = 688) were included in longitudinal outcomes analyses for a period of 90 days after hospital admission. Sankey diagrams were generated to show the patients’ transfers among the AD-strat clustering, liver transplant, ACLF development, death and survival status. The predictive power of the stratification models at different time points in the PREDICT cohort was evaluated using BIC, AIC, Concordance, and Likelihood ratio goodness-of-fit parameters (41).

## Discussion

Traditional patient classification methods based on outcome prediction scores may overlook important heterogeneity factors. To address this issue, we developed ClustALL, a computational pipeline that handles various clinical data challenges and identifies robust patient stratifications. We applied ClustALL to a cohort of patients with acute decompensation of cirrhosis, identifying five robust stratifications with prognostic value.

Optimal patient stratification is required to sustain the precision medicine revolution occurring in the clinical setting (43). While significant progress has been made in classification problems, particularly in domains like single-cell transcriptomic analysis (44,45), unsupervised clustering of patients based on clinical information is still in the developmental stage (7,46). Notably, the existing challenges in clinical stratification are often addressed using ad-hoc solutions that consider mixed data types, missing values, or highly correlated variables. However, no comprehensive method currently exists that addresses all these challenges. To overcome the aforementioned idiosyncrasies in clinical data, we have developed a novel computational framework named ClustALL. Significantly, beyond addressing existing challenges, ClustALL improves over other existing methodologies by allowing the identification of more than one robust stratification within a given population. Clinical data is complex and allows for multiple uses and “multiple interpretations” that may result in several valid groupings (47). Indeed, the concept of “multiple interpretations” arises from how variables are utilized in the clustering process and has been a subject of research in the early 21st century (48). Another distinctive feature of ClustALL is the consistency of the resulting representative stratifications even when limited modifications in the clustering parameter settings are applied. In the context of biological data, such as gene expression data, this property has already been already defined as the “propensity of a clustering algorithm to maintain output coherence over a range of settings”(49). Interestingly, this definition has been applied in the study of exposome and pregnancy-related mortality in the United States (50). In summary, we believe that ClustALL represents one of the initial necessary steps towards incorporating two necessary features into clinical stratification: parameter-based robustness and the identification of more than one stratification.

To assess the effectiveness of ClustALL, we applied it as a proof-of-concept in a cohort of patients with acutely decompensated cirrhosis considering data collected at hospital admission. Such an attempt to apply stratification to patients with cirrhosis has never been conducted. The stratification we set up differs from the scores developed and routinely used in patients with cirrhosis (e.g., MELD, MELD-Na, Child-Pugh, CLIF-C-AD) both in terms of design and use. Indeed, all these scores were built using a follow-up endpoint (usually death) in patients receiving therapies. These scores are helpful to identify patients at high risk of poor outcomes, but they do not fully capture the heterogeneity of the patients at admission for several reasons: (a) some features explaining patients heterogeneity might not have an independent prognostic value, either because the prognostic information they carry is contained in other variables, or because therapies administered to patients during their follow-up blunt their impact; (b) a similar survival rate does not imply similar pathophysiological mechanisms. For instance, in PREDICT, clusters 2 and 3 have a similar rate of ACLF, while they strongly differ with regard to the prevalence of hepatic encephalopathy. The stratification presented here is not intended to guide clinical bedside decisions or to replace a prognostic score but rather to identify homogeneous patient populations at hospital admission. This stratification could base the development of future clinical trials including more homogeneous patient populations. In this regard, to make this stratification easily accessible to all, we developed an online calculator and application available at https://decision-for-liver.eu/for-scientists/clustall-web-application/. The purely data-driven approach and the development and independent validation of the stratifications in large prospective multicentre European and Latin American cohorts strengthen our results.

Through this analysis, as a first step, we identified five alternative stratifications for patients with acute decompensation of cirrhosis. Interestingly, all stratifications included markers of impaired liver function, namely serum bilirubin and INR, but also the number of organ dysfunction or failure, and all but one included precipitating events. This emphasizes that these features are crucial when designing a clinical trial including patients with acute decompensation of cirrhosis. On the contrary, some features like age, sex, BMI, cause of cirrhosis and lifestyle were present in no or only one stratification suggesting that these features are not key when designing a clinical trial. The stratification we selected (AD-strat) provided a more granular resolution by allowing the identification of three subgroups of patients. In this stratification, diabetes mellitus is taken into account. While it is known that diabetes is an independent risk factor for cirrhosis decompensation (51,52), the role of diabetes once acute decompensation has happened has been overlooked so far. This place of diabetes is quite unique since causes of cirrhosis, comorbidities or lifestyle were not part of the key features of AD-strat. Hepatic encephalopathy strongly impacted the categorization of patients with acutely decompensated cirrhosis. Notably, 89% and 100% of the patients in Cluster 2 from the PREDICT and ACLARA cohorts, respectively, presented hepatic encephalopathy at the time of hospital admission. This may explain the intermediate prognosis observed in patients within Cluster 2, as hepatic encephalopathy is recognised by its fluctuating nature and potential reversibility (53,54). The dynamic nature of hepatic encephalopathy may also explain why Cluster 2 was not a static group over time (55).

Furthermore, tracking patients over time using AD-strat labelling allowed for dynamic and improved identification of patients at high risk of adverse events in the PREDICT cohort. These results highlight the ability of the ClustALL not only to stratify patients using baseline characteristics but also that the use of AD-strat labelling over time is able to improve this prediction.

Although our study showed promising results, it is important to acknowledge some limitations. Firstly, our stratification was based solely on routinely available clinical data at hospital admission, which may not provide a comprehensive view of patients’ conditions. Future studies should extend our findings with biological data, ideally derived from multiomic analyses. Moreover, it is crucial to consider that the predictive power in the ACLARA cohort was only assessed at 28 days due to the study design.

In conclusion, this study introduces a novel unsupervised clustering framework, ClustALL, able to overcome the limitations of previously available stratification methods. ClustALL is available as OpenSource (https://github.com/TranslationalBioinformaticsUnit/ClustALL_AD/). When applied to the setting of acute decompensation of cirrhosis, ClustALL enhanced our understanding of patients’ heterogeneity emphasizing the importance of liver function and the number of organ dysfunctions or failures, precipitating events, and conversely the limited role of age, sex, BMI, cause of cirrhosis and lifestyle at this stage of the liver disease. The selected stratification, AD-strat, might be a useful tool to better design future clinical trials by including more homogeneous patient populations.

## Data availability

Researchers who provide a methodology sound proposal can apply for the data, as far as the proposal is in line with the research consented by the patients. These proposals should be requested through https://www.clifresearch.com/decision/Home.aspx. Data requestors will need to sign a data transfer agreement. The code to generate the ClustALL method is available on GitHub, at https://github.com/TranslationalBioinformaticsUnit/ClustALL_AD/.

## Acknowledgements

This project has received funding from the European Union’s Horizon 2020 research and innovation program under grant agreement No 847949.

The study was supported by the European Foundation for the Study of Chronic Liver Failure (EF-Clif). The EF-Clif is a nonprofit private organization. The EF-Clif receives unrestricted donations from Cellex Foundation and Grifols. EF-Clif is partner, contributor and coordinator in several EU Horizon 2020 program projects. JT was appointed as visiting Professor in EF-Clif for the execution of the study by a grant from Cellex Foundation. The funders had no influence on study design, data collection and analysis, decision to publish or preparation of the manuscript.e fact that EF-CLIF.

Jonel Trebicka was supported by the German Research Foundation (DFG) project ID 403224013 – SFB 1382 (A09), by the German Federal Ministry of Education and Research (BMBF) for the DEEP-HCC project and by the Hessian Ministry of Higher Education, Research and the Arts (HMWK) for the ENABLE and ACLF-I cluster projects. The MICROB-PREDICT (project ID 825694), DECISION (project ID 847949), GALAXY (project ID 668031), LIVERHOPE (project ID 731875), and IHMCSA (project ID 964590) projects have received funding from the European Union’s Horizon 2020 research and innovation program. The manuscript reflects only the authors’ views, and the European Commission is not responsible for any use that may be made of the information it contains. The funders had no influence on study design, data collection and analysis, decision to publish, or preparation of the manuscript.

N.P.P was funded by a Ramón y Cajal fellow (RYC2021-032197-I) from the MCIN/AEI/10.13039/501100011033 and European Union “NextGenerationEU”/PRTR and by a Juan de la Cierva-formación fellow (FJC2019-042304-I) from the Spanish Ministry of Science and Innovation (MCIN).

P-E.R.’s research laboratory is supported by the Foundation pour la Recherche Médicale (FRM EQU202303016287), “Institut National de la Santé et de la Recherche Médicale” (ATIP AVENIR), the “Agence Nationale pour la Recherche” (ANR-18-CE14-0006-01, RHU QUID-NASH, ANR-18-IDEX-0001, ANR-22-CE14-0002) by « Émergence, Ville de Paris », by Fondation ARC and by the European Union’s Horizon 2020 research and innovation programme under grant agreement No 847949.

