## Supplementary Material for "ClustALL: A robust clustering strategy for stratification of patients with acutely decompensated cirrhosis"

**Figure S1. Schematic overview of the different steps of the ClustALL approach using the framework with missing values (best viewed in colour)**. First, (A) a dendrogram and its associated depths are computed considering the original dataset with missing values. The original dataset is then (B). imputed 1,000 times with the MICE algorithm. Then, (C) the 1,000 complete datasets for each resulting depth of the dendrogram are reduced using Principal Component Analysis (PCA). The output is an embedding for each possible depth and imputation (Green Panel). Then, stratification is computed considering the combination distance metric, clustering algorithm and each *Embedding* derived from an imputed dataset (purple panel A). The selection of the optimal number of clusters is based on the consensus from cluster internal validation and the mode of the imputed datasets for each corresponding embedding (purple panel B). Afterwards, a distance matrix ($D_{mat}$) between individuals is obtained by computing how often two individuals are assigned to the same cluster in each imputation Then, $D_{mat}$ is used to calculate a final stratification using correlation-based distance and h-clust (purple panel C). The final step follows the ClustALL framework without imputations (red panel).

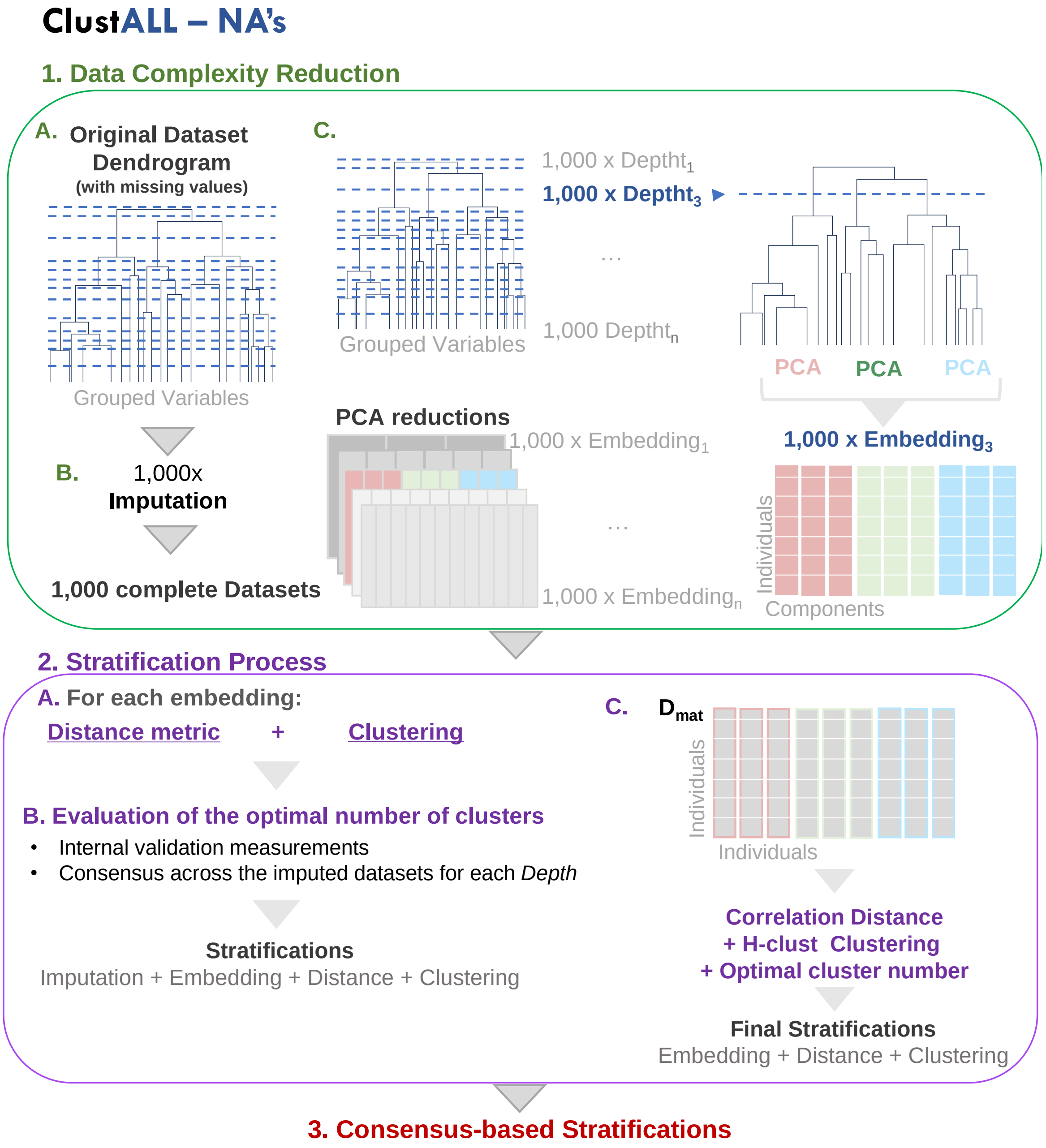

**Figure S2.** ***Principal Component projections of the ClustALL robust stratifications based on all the input variables.*** (A-E). Low-dimension representation of the robust stratifications after applying the ClustALL framework to the PREDICT cohort. For each one of the 5 robust stratifications identified by ClustALL, the Principal Component Analysis of all the input variables with the stratification is shown. The *x* (Dim1) and *y* (Dim2) axes represent the first and second principal components respectively, which are linear combinations of the original variables.

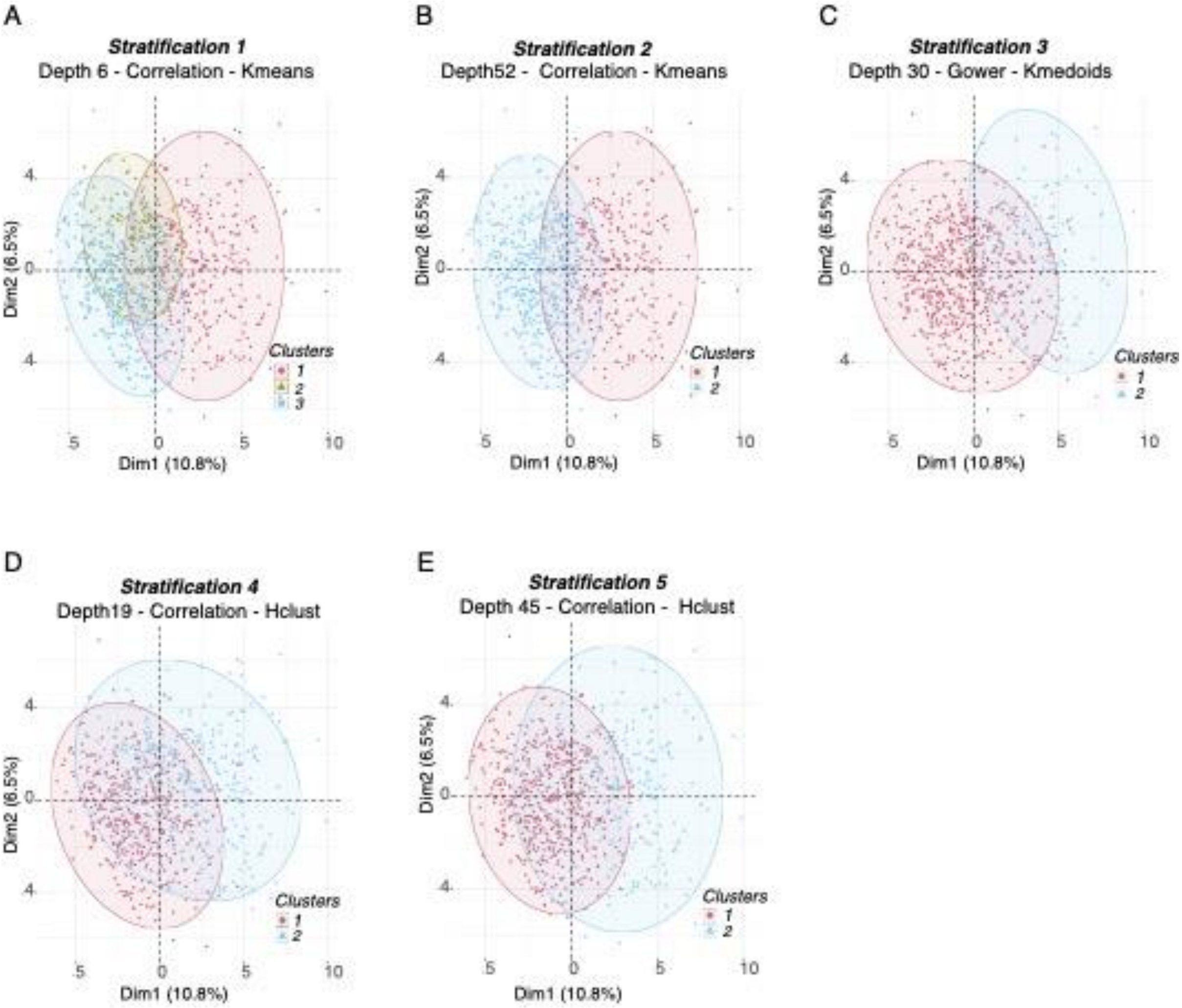

**Figure S3. Frequency Density Plot from bootstrapping the Predict cohort stratification**. The figure displays a bootstrapped density plot used for stratifying the PREDICT data at hospital admission computed within the traditional clustering methods (blue line) versus ClustALL framework (red line), 1,000 times. The x-axis represents the frequency, while the y-axis depicts the resampling times. Traditional methods consider k-means or hierarchical clustering method plus correlation or Gower distances, directly applied to data without considering any dimensionality reduction step.

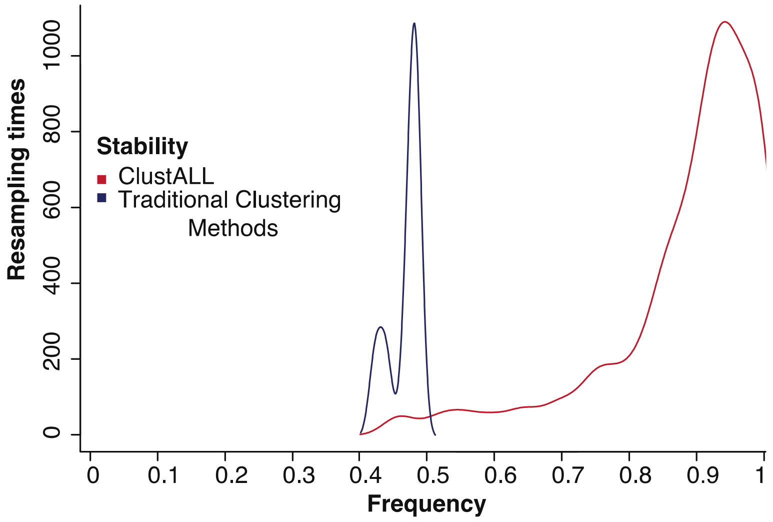


**Figure S4. Visual representation of the population-based robustness concept**. The figure displays a bootstrapped density plot used for stratifying the PREDICT data at hospital admission computed with ClustALL framework (red line), 1,000 times. The x-axis represents the frequency, while the y-axis depicts the resampling times. The discontinuous lines indicate that more than 85% of the stratifications remain stable despite resampling.

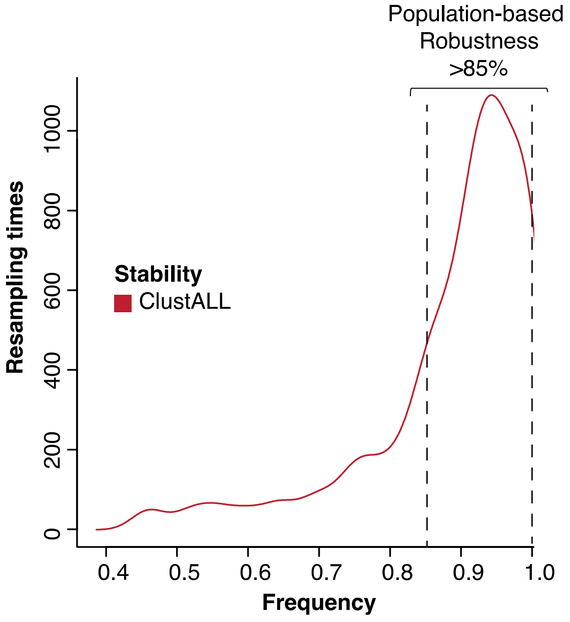

**Figure S5. The window of time between visit and reported event.** Distribution of the window of time (in days) between the reported event and hospital admission visit (in gray) or the reported event and the last reported visit (in blue). Patients with any reported event and at least one visit after hospital admission are included (147). The difference between the window of times is significant (Wilcoxon test p-value <0.001).

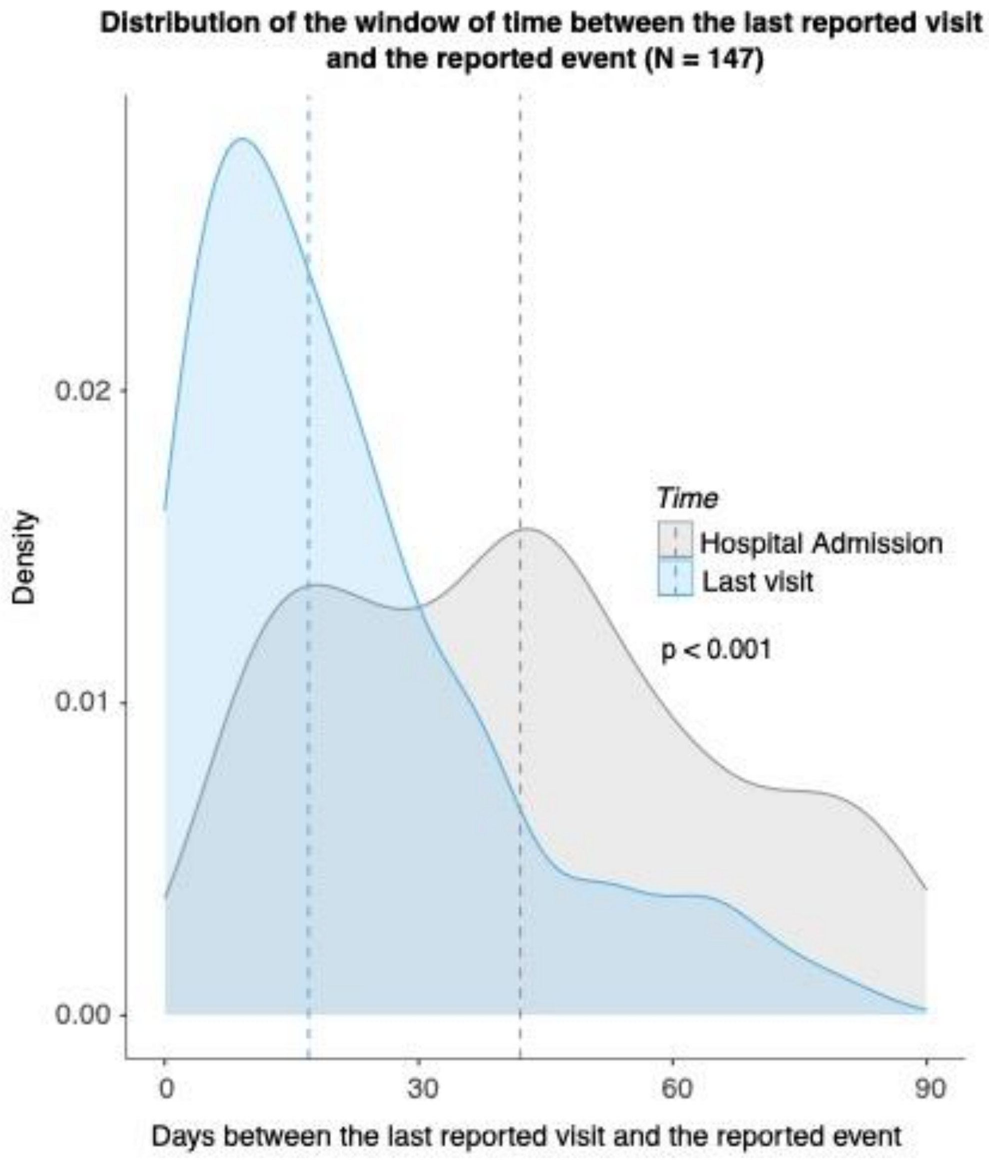

**Table S1. Descriptive statistics of the characteristics for 766 PREDICT patients collected at Hospital Admission**. The values on the left from 1 to 74 indicate the number of variables included as input in the ClustALL framework. The first column represents all the input variables. The second column represents the number and corresponding percentage of missing values per variable. The third to the sixth columns represent the number of patients per variable and corresponding percentage or mean values ± SD for each variable across all patients for the entire cohort, cluster 1, cluster 2 and cluster 3 respectively. The p-values are from the ANOVA and Chi-square analysis for continuous and categorical binary variables between the three clusters.

| | **Patient Characteristics (PREDICT)** | **Number of missing values** | **Overall population (n=766)** | **Cluster 1 (n=306)** | **Cluster 2 (n=118)** | **Cluster 3 (n=342)** | ***P-value*** |
|---|---|---|---|---|---|---|---|
| | **Demographics** | | | | | | |
| (1) | Age — years | 0 (0) | 59.44 ± 10.81 | 56.81 ± 10.01 | 62.88 ± 11.6 | 60.61 ± 10.7 | <0.01 |
| (2) | Sex — no.(%) | 0 (0) | 247 (32.25) | 103 (33.66) | 31 (26.27) | 113 (33.04) | 0.3 |
| (3) | Height — cm | 0 (0) | 170.07 ± 9.21 | 170.54 ± 9.23 | 169.65 ± 9.2 | 169.8 ± 9.2 | 0.31 |
| (4) | Weight — kg | 14 (1.83) | 78.4 ± 16.42 | 78.66 ± 17 | 77.53 ± 15.55 | 78.46 ± 16.23 | 0.89 |
| (5) | Body Mass Index (kg/m²) | 14 (1.83) | 27.05 ± 5.01 | 26.96 ± 4.98 | 26.95 ± 5.19 | 27.17 ± 4.99 | 0.58 |
| (6) | Ethnicity Black or African American — no.(%) | 0 (0) | 2 (0.26) | 0 (0) | 2 (1.69) | 0 (0) | <0.01 |
| (7) | Ethnicity Asian — no.(%) | 0 (0) | 12 (1.57) | 8 (2.61) | 4 (3.39) | 0 (0) | <0.01 |
| (8) | Ethnicity White — no.(%) | 0 (0) | 740 (96.61) | 295 (96.41) | 103 (87.29) | 342 (100) | <0.01 |
| (9) | Ethnicity Other — no.(%) | 0 (0) | 12 (1.57) | 3 (0.98) | 9 (7.63) | 0 (0) | <0.01 |
| | **Cause of cirrhosis** | | | | | | |
| (10) | Etiology Alcohol — no.(%) | 0 (0) | 454 (59.27) | 182 (59.48) | 70 (59.32) | 202 (59.06) | 0.99 |
| (11) | Etiology Viral — no.(%) | 0 (0) | 83 (10.84) | 32 (10.46) | 20 (16.95) | 31 (9.06) | 0.06 |
| (12) | Etiology Alcohol + Viral — no.(%) | 0 (0) | 66 (8.62) | 36 (11.76) | 6 (5.08) | 24 (7.02) | 0.03 |
| (13) | Etiology NASH — no.(%) | 0 (0) | 65 (8.49) | 20 (6.54) | 10 (8.47) | 35 (10.23) | 0.24 |
| (14) | Etiology Other — no.(%) | 0 (0) | 61 (7.96) | 22 (7.19) | 10 (8.47) | 29 (8.48) | 0.81 |
| (15) | Etiology Cryptogenic— no.(%) | 0 (0) | 37 (4.83) | 14 (4.58) | 2 (1.69) | 21 (6.14) | 0.15 |
| | **Main reason for hospitalization** | | | | | | |
| (16) | Ascites — no.(%) | 0 (0) | 330 (43.08) | 115 (37.58) | 21 (17.8) | 194 (56.73) | <0.01 |
| (17) | Hepatic encephalopathy — no.(%) | 0 (0) | 120 (15.67) | 48 (15.69) | 69 (58.47) | 3 (0.88) | <0.01 |
| (18) | Gastrointestinal bleeding — no.(%) | 0 (0) | 116 (15.14) | 24 (7.84) | 16 (13.56) | 76 (22.22) | <0.01 |
| (19) | Spontaneous bacterial peritonitis — no.(%) | 0 (0) | 22 (2.87) | 17 (5.56) | 0 (0) | 5 (1.46) | <0.01 |
| (20) | Other infection — no.(%) | 0 (0) | 79 (10.31) | 47 (15.36) | 6 (5.08) | 26 (7.6) | <0.01 |
| | **Manifestations at admission** | | | | | | |
| (21) | Ascites — no.(%) | 0 (0) | 542 (70.76) | 238 (77.78) | 52 (44.07) | 252 (73.68) | <0.01 |
| (22) | Hepatic encephalopathy — no.(%) | 0 (0) | 234 (30.55) | 127 (41.5) | 105 (88.98) | 2 (0.58) | <0.01 |
| (23) | Gastrointestinal bleeding — no.(%) | 0 (0) | 108 (14.1) | 24 (7.84) | 19 (16.1) | 65 (19.01) | <0.01 |

| | | | | | | | |
|---|---|---|---|---|---|---|---|
| (24) | Acute kidney injury — no.(%) | 0 (0) | 45 (5.87) | 27 (8.82) | 4 (3.39) | 14 (4.09) | 0.02 |
| (25) | Bacterial Infection — no.(%) | 0 (0) | 222 (28.98) | 139 (45.42) | 23 (19.49) | 60 (17.54) | <0.01 |
| (26) | Alcoholic Hepatitis — no.(%) | 0 (0) | 55 (7.18) | 47 (15.36) | 0 (0) | 8 (2.34) | <0.01 |
| (27) | Acute Viral Hepatitis — no.(%) | 0 (0) | 5 (0.65) | 3 (0.98) | 0 (0) | 2 (0.58) | 0.52 |
| (28) | Hepatocellular Carcinoma — no.(%) | 0 (0) | 34 (4.44) | 12 (3.92) | 6 (5.08) | 16 (4.68) | 0.84 |
| (29) | Number of clinical events — number | 0 (0) | 1.63 ± 0.85 | 2.02 ± 0.9 | 1.77 ± 0.8 | 1.23 ± 0.59 | <0.01 |
| (30) | Number of precipitating events — number | 0 (0) | 0.27 ± 0.49 | 0.48 ± 0.59 | 0.12 ± 0.32 | 0.13 ± 0.34 | <0.01 |
| (31) | Liver Dysfunction— no.(%) * | 0 (0) | 121 (15.8) | 109 (35.62) | 2 (1.69) | 10 (2.92) | <0.01 |
| (32) | Renal Dysfunction — no.(%) * | 0 (0) | 63 (8.22) | 27 (8.82) | 9 (7.63) | 27 (7.89) | 0.88 |
| (33) | Brain Dysfunction — no.(%) * | 0 (0) | 215 (28.07) | 119 (38.89) | 94 (79.66) | 2 (0.58) | <0.01 |
| (34) | Coagulation Dysfunction — no.(%) * | 0 (0) | 61 (7.96) | 61 (19.93) | 0 (0) | 0 (0) | <0.01 |
| (35) | Heart Dysfunction — no.(%) * | 0 (0) | 83 (10.84) | 47 (15.36) | 5 (4.24) | 31 (9.06) | <0.01 |
| (36) | Respiratory Dysfunction — no.(%) * | 0 (0) | 31 (4.05) | 20 (6.54) | 2 (1.69) | 9 (2.63) | <0.01 |
| (37) | Liver Failure — no.(%) * | 0 (0) | 36 (4.7) | 36 (11.76) | 0 (0) | 0 (0) | <0.01 |
| (38) | Brain Liver Failure — no.(%) * | 0 (0) | 19 (2.48) | 8 (2.61) | 11 (9.32) | 0 (0) | <0.01 |
| (39) | Coagulation Failure — no.(%) * | 0 (0) | 17 (2.22) | 16 (5.23) | 0 (0) | 1 (0.29) | <0.01 |
| (40) | Heart Failure — no.(%) * | 0 (0) | 3 (0.39) | 2 (0.65) | 0 (0) | 1 (0.29) | 0.58 |
| (41) | Respiratory Failure — no.(%) * | 0 (0) | 1 (0.13) | 0 (0) | 0 (0) | 1 (0.29) | 0.54 |
| (42) | Number of dysfunctions — number | 0 (0) | 0.75 ± 0.86 | 1.25 ± 0.98 | 0.95 ± 0.54 | 0.23 ± 0.45 | <0.01 |
| (43) | Number of failures — number | 0 (0) | 0.1 ± 0.3 | 0.2 ± 0.4 | 0.09 ± 0.29 | 0.01 ± 0.09 | <0.01 |
| | **Cirrhosis Severity Scores** | | | | | | |
| (44) | CLIF-C-AD | 62 (8.09) | 51.89 ± 7.95 | 56.99 ± 6.58 | 48.53 ± 6.4 | 48.48 ± 7.08 | <0.01 |
| (45) | MELD-NA | 0 (0) | 18.61 ± 5.48 | 23.3 ± 3.78 | 15.61 ± 3.31 | 15.46 ± 4.25 | <0.01 |
| (46) | MELD | 0 (0) | 15.65 ± 5.01 | 19.92 ± 3.93 | 13.29 ± 2.74 | 12.65 ± 3.57 | <0.01 |
| (47) | Child-Pugh | 0 (0) | 2.11 ± 0.58 | 2.54 ± 0.52 | 1.92 ± 0.32 | 1.8 ± 0.45 | <0.01 |
| (48) | CLIF-OF | 0 (0) | 6.87 ± 0.98 | 7.57 ± 1.04 | 7.06 ± 0.48 | 6.17 ± 0.41 | <0.01 |
| | **Medical History** | | | | | | |
| (49) | Diabetes Mellitus — no.(%) | 0 (0) | 222 (28.98) | 47 (15.36) | 60 (50.85) | 115 (33.63) | <0.01 |
| (50) | Arterial hypertension — no.(%) | 0 (0) | 264 (34.46) | 89 (29.08) | 48 (40.68) | 127 (37.13) | 0.03 |
| (51) | History of previous decompensations — no.(%) | 13 (1.7) | 631 (82.38) | 239 (78.1) | 102 (86.44) | 290 (84.8) | 0.04 |
| | **Lifestyle** | | | | | | |
| (52) | Alcohol consumption in the 3 previous months — no.(%) | 125 (16.32) | 295 (38.51) | 147 (48.04) | 21 (17.8) | 127 (37.13) | <0.01 |
| (53) | Active alcohol consumption — no.(%) | 130 (16.97) | 216 (28.19) | 113 (14.75) | 14 (18.27) | 89 (11.61) | <0.01 |
| (54) | Active smoker — no.(%) | 2 (0.26) | 423 (55.22) | 167 (54.58) | 52 (44.07) | 204 (59.65) | 0.01 |
| | **Laboratory Variables** | | | | | | |

| | | | | | | | |
|---|---|---|---|---|---|---|---|
| (55) | Serum Alanine Transaminase — U/L | 17 (2.22) | 37.85 ± 42.4 | 50.81 ± 60.01 | 28.65 ± 19.24 | 29.42 ± 20.83 | <0.01 |
| (56) | Serum Aspartate aminotransferase — U/L | 52 (6.79) | 70.38 ± 72.32 | 96.41 ± 93.01 | 47.66 ± 29.42 | 54.94 ± 51.7 | <0.01 |
| (57) | Serum Albumin — g/dL | 45 (5.87) | 2.9 ± 0.59 | 2.7 ± 0.52 | 2.96 ± 0.56 | 3.06 ± 0.61 | <0.01 |
| (58) | Serum Total Bilirubin — mg/dL | 0 (0) | 4.05 ± 4.36 | 7.02 ± 5.4 | 2.13 ± 1.49 | 2.06 ± 1.62 | <0.01 |
| (59) | Serum Gamma-glutamyl transferase — U/L | 94 (12.27) | 170.57 ± 244.25 | 182.6 ± 232.68 | 92.91 ± 96.48 | 186.6 ± 282.38 | 0.77 |
| (60) | Serum C-Reactive Protein — mg/L | 85 (11.1) | 27.28 ± 35.09 | 37.83 ± 38.85 | 15.96 ± 23.08 | 21.74 ± 32.47 | <0.01 |
| (61) | Serum Sodium— mEq/L | 0 (0) | 135.26 ± 5.28 | 133.33 ± 5.5 | 137.02 ± 4.51 | 136.39 ± 4.77 | <0.01 |
| (62) | Serum Potassium — mEq/L | 8 (1.04) | 4.04 ± 0.64 | 4 ± 0.72 | 4.06 ± 0.56 | 4.08 ± 0.59 | 0.13 |
| (63) | Serum Glucose — mg/dL | 132 (17.23) | 124.03 ± 51.33 | 119.04 ± 44.33 | 136.51 ± 54.15 | 124.19 ± 55.41 | 0.23 |
| (64) | Hemoglobin — g/dL | 1 (0.13) | 10.39 ± 2.23 | 10.42 ± 2.2 | 10.5 ± 2.11 | 10.32 ± 2.29 | 0.53 |
| (65) | Hematocrit — % | 14 (1.83) | 30.76 ± 6.17 | 30.43 ± 6.14 | 31.16 ± 5.95 | 30.91 ± 6.27 | 0.33 |
| (66) | Creatinine — mg/dL | 0 (0) | 0.95 ± 0.34 | 0.94 ± 0.36 | 0.99 ± 0.35 | 0.96 ± 0.33 | 0.45 |
| (67) | White blood cell count — x10^9/L | 0 (0) | 6.97 ± 4.08 | 8.87 ± 4.72 | 5.09 ± 2.06 | 5.91 ± 3.22 | <0.01 |
| (68) | Lymphocytes — x10^9/L | 145 (18.93) | 1.13 ± 0.61 | 1.26 ± 0.65 | 1.09 ± 0.63 | 1.02 ± 0.55 | <0.01 |
| (69) | Monocytes— x10^9/L | 157 (20.5) | 0.67 ± 0.38 | 0.84 ± 0.41 | 0.54 ± 0.29 | 0.56 ± 0.3 | <0.01 |
| (70) | Neutrophils— x10^9/L | 133 (17.36) | 4.76 ± 3.46 | 6.29 ± 4.13 | 3.16 ± 1.58 | 3.95 ± 2.66 | <0.01 |
| (71) | INR (International Normalized Ratio) | 0 (0) | 1.52 ± 0.48 | 1.79 ± 0.63 | 1.36 ± 0.19 | 1.34 ± 0.22 | <0.01 |
| (72) | Platelet — x10^3/µL | 7 (0.91) | 115.22 ± 75.35 | 114.89 ± 80.93 | 99.14 ± 60.59 | 121.06 ± 74.14 | 0.27 |
| (73) | SpO2 — (%) | 48 (6.27) | 97 ± 2.12 | 96.82 ± 2.27 | 97 ± 2.17 | 97.17 ± 1.95 | 0.04 |
| (74) | SpO2/FiO2 Ratio | 48 (6.27) | 456.27 ± 30.66 | 452.45 ± 37.31 | 459.07 ± 25.23 | 458.71 ± 24.97 | 0.01 |
| | **Outcomes** | | | | | | |
| | 28-day mortality — no.(%) | 0 (0) | 34 (4.44) | 23 (3) | 3 (0.39) | 8 (1.04) | <0.01 |
| | 90-day mortality — no.(%) | 0 (0) | 103 (13.45) | 62 (8.09) | 5 (0.65) | 36 (4.70) | <0.01 |
| | 28-day ACLF development — no.(%) | 0 (0) | 64 (8.36) | 43 (5.61) | 6 (0.78) | 15 (1.96) | <0.01 |
| | 90-day ACLF development — no.(%) | 0 (0) | 114 (14.88) | 70 (9.14) | 10 (1.31) | 34 (4.44) | <0.01 |
| | 28-day transplant — no.(%) | 0 (0) | 17 (2.22) | 13 (1.70) | 1 (0.13) | 3 (0.39) | <0.01 |
| | 90-day transplant — no.(%) | 0 (0) | 49 (6.40) | 35 (4.57) | 2 (0.26) | 12 (1.57) | <0.01 |

*ACLF=Acute-on-chronic liver failure, FiO2= Fraction of inspired oxygen NASH= Non-alcoholic steatohepatitis, SpO2= Saturation of peripheral Oxygen.*

** as defined in Jalan, R., et al. (2014). Development and validation of a prognostic score to predict mortality in patients with acute-on-chronic liver failure. Journal of Hepatology, 61(5), 1038–1047. https://doi.org/10.1016/j.jhep.2014.06.012*

**Table S2. Patient distribution through the clusters when using the classical clustering methodologies**. The distribution of the PREDICT patients after applying different distance metrics (Correlation and Gower) and clustering techniques (K-means, Hierarchical Clustering, and K-medoids) considering 2 or 3 clusters are shown.

| ***Correlation K-means*** | | | |
|---|---|---|---|
| ***k=2*** | *Cluster 1* = 734 | *Cluster 2* = 32 | |
| ***k=3*** | *Cluster 1* = 732 | *Cluster 2* = 25 | *Cluster 3* = 9 |

| ***Correlation Hierarchical Clustering*** | | | |
|---|---|---|---|
| ***k=2*** | *Cluster 1* = 757 | *Cluster 2* = 9 | |
| ***k=3*** | *Cluster 1* = 734 | *Cluster 2* = 23 | *Cluster 3* = 9 |

| ***Gower K-medoids*** | | | |
|---|---|---|---|
| ***k=2*** | *Cluster 1* = 512 | *Cluster 2* = 254 | |
| ***k=3*** | *Cluster 1* = 321 | *Cluster 2* = 232 | *Cluster 3* = 213 |

| ***Gower Hierarchical Clustering*** | | | |
|---|---|---|---|
| ***k=2*** | *Cluster 1* = 655 | *Cluster 2* = 111 | |
| ***k=3*** | *Cluster 1* = 360 | *Cluster 2* = 295 | *Cluster 3* = 111 |

**Table S3-S7. Summary of the *most predictive* variables in the identified clusters for 1 to 5 Stratification representatives in PREDICT cohort at *hospital admission*.** The p-values are derived from the ANOVA analysis for continuous variables or from $\chi 2$ analysis for binary and categorical variables. Values are the mean ± SD unless indicated otherwise.

**Table S3. Stratification 1** *(Height 6, Correlation, K-means).*

| Clinical Feature | Cluster 1 (n=306) | Cluster 2 (n=118) | Cluster 3 (n=342) | *P-value* |
|---|---|---|---|---|
| Diabetes Mellitus — no.(%) | 47 (15.36) | 60 (50.85) | 115 (33.63) | 1.90E-13 |
| Hepatic Encephalopathy — no.(%) | 127 (41.5) | 105 (88.98) | 2 (0.58) | 3.77E-77 |
| Number of clinical events — number | 2.02 ± 0.9 | 1.77 ± 0.8 | 1.23 ± 0.59 | 1.17495E-35 |
| Organ Dysfunction — number | 1.25 ± 0.98 | 0.95 ± 0.54 | 0.23 ± 0.45 | 9.42939E-61 |
| Number of precipitating events — number | 0.48 ± 0.59 | 0.12 ± 0.32 | 0.13 ± 0.34 | 1.04935E-22 |
| Serum Albumin — g/dL | 2.7 ± 0.52 | 2.96 ± 0.56 | 3.06 ± 0.61 | 6.64445E-15 |
| Serum Aspartate Aminotransferase — units/L | 96.41 ± 93.01 | 47.66 ± 29.42 | 54.94 ± 51.7 | 6.87725E-16 |
| Serum CRP — mg/L | 37.83 ± 38.85 | 15.96 ± 23.08 | 21.74 ± 32.47 | 1.46111E-11 |
| Serum Bilirubin —mg/dL | 7.02 ± 5.4 | 2.13 ± 1.49 | 2.06 ± 1.62 | 7.86805E-62 |
| INR | 1.79 ± 0.63 | 1.36 ± 0.19 | 1.34 ± 0.22 | 5.42777E-39 |
| Serum Sodium — mEq/L | 133.33 ± 5.5 | 137.02 ± 4.51 | 136.39 ± 4.77 | 1.6674E-16 |
| White Blood Cells — x$10^3$/$mm^3$ | 8.87 ± 4.72 | 5.09 ± 2.06 | 5.91 ± 3.22 | 1.27361E-27 |

**CRP=C-Reactive Protein, INR= International Normalized Ratio.*

¶ *Counting the hospital admission visit.*

**Table S4.** Stratification 2 *(Height 30, Correlation, K-means).*

| Clinical Feature | Cluster 1 (n=283) | Cluster 2 (n=483) | *P-value* |
|---|---|---|---|
| Diabetes Mellitus — no.(%) | 45 (15.9) | 177 (36.65) | 1.68E-09 |
| Gastrointestinal Bleeding — no.(%) | 20 (7.07) | 88 (18.22) | 3.00E-05 |
| Number of clinical events — number | 2.04 ± 0.9 | 1.38 ± 0.71 | 1.84E-23 |
| Respiratory Dysfunction — no.(%) | 23 (8.13) | 8 (1.66) | 2.71E-05 |
| Organ Dysfunction — number | 1.28 ± 0.99 | 0.44 ± 0.59 | 3.58E-36 |
| Number of precipitating events — number | 0.51 ± 0.6 | 0.13 ± 0.33 | 3.84E-24 |
| Serum Albumin — g/dL | 2.7 ± 0.52 | 3.02 ± 0.6 | 5.29E-13 |
| CRP — mg/L | 39.2 ± 39.7 | 20.3 ± 30 | 2.58E-13 |
| Serum Bilirubin—mg/dL | 7.33 ± 5.5 | 2.13 ± 1.58 | 1.04E-68 |
| INR | 1.8 ± 0.65 | 1.36 ± 0.23 | 1.01E-38 |
| Serum Sodium — mEq/L | 133.2 ± 5.44 | 136.47 ± 4.79 | 2.16E-17 |
| White Blood Cells — x$10^3$/$mm^3$ | 8.93 ± 4.79 | 5.82 ± 3.07 | 4.43E-26 |

**CRP=C-Reactive Protein, INR= International Normalized Ratio.*

¶ *Counting the hospital admission visit.*

**Table S5.** Stratification 3 (*Height 30, Gower, H-clust).*

| Clinical Feature | Cluster 1 (n=666) | Cluster 2 (n=100) | *P-value* |
|---|---|---|---|
| Number of clinical events — number | 1.53 ± 0.78 | 2.27 ± 0.99 | 6.47E-17 |
| Liver Dysfunction — no.(%) | 59 (8.86) | 62 (62) | 3.55E-41 |
| Cerebral Dysfunction — no.(%) | 168 (25.23) | 47 (47) | 1.09E-05 |
| Coagulation Dysfunction — no.(%) | 0 (0) | 61 (61) | 3.39E-96 |
| Number of dysfunctions — number | 0.56 ± 0.66 | 1.99 ± 1.02 | 2.68E-60 |
| Liver Failure — no.(%) | 23 (3.45) | 13 (13) | 7.72E-05 |
| INR | 1.44 ± 0.32 | 2.08 ± 0.86 | 2.41E-39 |
| Serum Sodium — mEq/L | 135.59 ± 5.1 | 133.1 ± 5.92 | 1.01E-05 |

**INR= International Normalized Ratio .*

¶ *Counting the hospital admission visit.*

**Table S6.** Stratification 4 (*Height* 19, Correlation, *H-clust*)

| Clinical Feature | Cluster 1 (n=431) | Cluster 2 (n=335) | *P-value* |
|---|---|---|---|
| Age — number | 60.19 ± 10.6 | 58.48 ± 11.01 | 3.00E-02 |
| Hepatic encephalopathy — no.(%) | 34 (7.89) | 200 (59.7) | 2.83E-53 |
| Gastrointestinal Bleeding — no.(%) | 81 (18.79) | 27 (8.06) | 3.63E-05 |
| Acute Viral Hepatitis — no.(%) | 5 (1.16) | 0 (0) | 1.27E-01 |
| Liver Dysfunction — no.(%) | 30 (6.96) | 91 (27.16) | 6.11E-14 |
| Coagulation Dysfunction — no.(%) | 2 (0.46) | 59 (17.61) | 1.11E-17 |
| Respiratory Dysfunction — no.(%) | 0 (0) | 31 (9.25) | 3.79E-10 |
| Number of failures — number | 0 ± 0.05 | 0.22 ± 0.42 | 8.89E-24 |

¶ *Counting the hospital admission visit.*

**Table S7.** *Stratification 5* (*Height* 45, Correlation, *H-clust*).

| Clinical Feature | Cluster 1 (n=522) | Cluster 2 (n=244) | *P-value* |
|---|---|---|---|
| Alcohol etiology — no.(%) | 295 (56.51) | 159 (65.16) | 2.84E-02 |
| Gastrointestinal Bleeding — no.(%) | 94 (18.01) | 14 (5.74) | 9.21E-06 |
| Acute Alcoholic-steatohepatitis — no.(%) | 6 (1.15) | 49 (20.08) | 1.32E-20 |
| Hepatocellular carcinoma — no.(%) | 29 (5.56) | 5 (2.05) | 4.47E-02 |
| Liver Dysfunction — no.(%) | 25 (4.79) | 96 (39.34) | 9.21E-34 |
| Coagulation Dysfunction — no.(%) | 3 (0.57) | 58 (23.77) | 1.09E-27 |
| Respiratory Dysfunction — no.(%) | 4 (0.77) | 27 (11.07) | 6.04E-11 |
| Number of failures — number | 0.01 ± 0.09 | 0.3 ± 0.46 | 1.35E-34 |
| Number of precipitating events — number | 0.17 ± 0.38 | 0.48 ± 0.6 | 2.18E-16 |
| Serum Bilirubin —mg/dl | 2.46 ± 1.91 | 7.45 ± 5.92 | 1.14E-57 |
| Serum Sodium — mEq/L | 136.04 ± 5.04 | 133.6 ± 5.39 | 1.54E-09 |
| Smoker — no.(%) | 304 (58.24) | 119 (48.77) | 1.75E-02 |

¶ *Counting the inclusion visit.*

**Table S8.** Distribution of the *AD-strat* clustering-based labelling in PREDICT cohort at hospital admission obtained with ClustALL vs. acute decompensation of cirrhosis classification as defined in The PREDICT study from Trebicka, J, et al.

| N =766 | SDC | UDC | pre-ACLF |
|---|---|---|---|
| **Cluster 1 — no.(%)** | 157 (33.7) | 78 (42.4) | 71 (61.2) |
| **Cluster 2 — no.(%)** | 83 (17.8) | 25 (13.6) | 10 (8.6) |
| **Cluster 3 — no.(%)** | 226 (48.5) | 81 (44) | 35 (30.2) |

**pre-ACLF= pre-acute-on-chronic liver failure, SDC=Stable decompensated cirrhosis, UDC= Unstable decompensated cirrhosis.*

**Table S9. Characteristics for 580 ACLARA patients collected at Hospital Admission**. The first column represents the ACLARA variables. The second column represents the number and corresponding percentage of missing values per variable. The third to the sixth columns represent the number of patients per variable and corresponding percentage or mean values ± SD for each variable across all patients for the entire cohort, cluster 1, cluster 2 and cluster 3 respectively. The p-values are from the ANOVA and Chi-square analysis for continuous and categorical binary variables between the three clusters.

| Patient Characteristics (ACLARA) | Number of missing values | Overall population (n=580) | Cluster 1 (n=185) | Cluster 2 (n=99) | Cluster 3 (n=296) | *P-value* |
|---|---|---|---|---|---|---|
| **Demographics** | | | | | | |
| Age — years | 0 (0) | 58.1 ± 12.3 | 54.08 ± 12.44 | 62.74 ± 10.04 | 59.06 ± 12.19 | <0.01 |
| Sex — no.(%) | 0 (0) | 205 (35.34) | 50 (27.03) | 33 (33.33) | 122 (41.22) | <0.01 |
| Body Mass Index (kg/m²) | 103 (17.76) | 27.11 ± 4.76 | 27.21 ± 4.93 | 27.26 ± 4.67 | 27.01 ± 4.7 | 0.63 |
| Ethnicity Black or African American — no.(%) | 0 (0) | 105 (18.1) | 38 (20.54) | 21 (21.21) | 46 (15.54) | 0.26 |
| Ethnicity Asian — no.(%) | 0 (0) | 1 (0.17) | 0 (0) | 0 (0) | 1 (0.34) | 0.62 |
| Ethnicity Indian or Alaska Native American— no.(%) | 0 (0) | 98 (16.9) | 40 (21.62) | 15 (15.15) | 43 (14.53) | 0.11 |
| Ethnicity White — no.(%) | 0 (0) | 301 (51.9) | 82 (44.32) | 53 (53.54) | 166 (56.08) | 0.12 |
| Multiple — no.(%) | 0 (0) | 75 (12.93) | 25 (13.51) | 10 (10.1) | 40 (13.51) | 0.65 |
| **Cause of cirrhosis** | | | | | | |
| Etiology Alcohol — no.(%) | 0 (0) | 208 (35.86) | 60 (32.43) | 36 (36.36) | 112 (37.84) | 0.48 |
| Etiology Viral — no.(%) | 0 (0) | 77 (13.28) | 23 (12.43) | 14 (14.14) | 40 (13.51) | 0.91 |
| Etiology Alcohol + Viral — no.(%) | 0 (0) | 35 (6.03) | 10 (5.41) | 5 (5.05) | 20 (6.76) | 0.75 |
| Etiology NASH — no.(%) | 0 (0) | 102 (17.59) | 35 (18.92) | 18 (18.18) | 49 (16.55) | 0.79 |
| Etiology Other — no.(%) | 0 (0) | 66 (11.38) | 18 (9.73) | 12 (12.12) | 36 (12.16) | 0.69 |
| Etiology Cryptogenic— no.(%) | 0 (0) | 92 (15.86) | 39 (21.08) | 14 (14.14) | 39 (13.18) | 0.06 |
| **Main reason for hospitalization** | | | | | | |
| Ascites — no.(%) | 0 (0) | 138 (23.79) | 40 (21.62) | 12 (12.12) | 86 (29.05) | <0.01 |
| Hepatic encephalopathy — no.(%) | 0 (0) | 109 (18.79) | 50 (27.03) | 53 (53.54) | 6 (2.03) | <0.01 |
| Gastrointestinal bleeding — no.(%) | 0 (0) | 175 (30.17) | 28 (15.14) | 20 (20.2) | 127 (42.91) | <0.01 |
| Spontaneous bacterial peritonitis — no.(%) | 0 (0) | 51 (8.79) | 24 (12.97) | 3 (3.03) | 24 (8.11) | 0.02 |
| Other infection — no.(%) | 0 (0) | 83 (14.31) | 32 (17.3) | 8 (8.08) | 43 (14.53) | 0.11 |
| **Manifestations at admission** | | | | | | |
| Ascites — no.(%) | 0 (0) | 365 (62.93) | 145 (78.38) | 48 (48.48) | 172 (58.11) | <0.01 |
| Hepatic encephalopathy — no.(%) | 0 (0) | 189 (32.59) | 90 (48.65) | 99 (100) | 0 (0) | <0.01 |
| Gastrointestinal bleeding — no.(%) | 0 (0) | 183 (31.55) | 33 (17.84) | 21 (21.21) | 129 (43.58) | <0.01 |
| Acute kidney injury — no.(%) | 0 (0) | 87 (15) | 37 (20) | 8 (8.08) | 42 (14.19) | 0.02 |
| Bacterial Infection — no.(%) | 0 (0) | 242 (41.72) | 108 (58.38) | 33 (33.33) | 101 (34.12) | <0.01 |
| Alcoholic Hepatitis — no.(%) | 0 (0) | 122 (21.03) | 75 (40.54) | 11 (11.11) | 36 (12.16) | <0.01 |

| | | | | | | |
|---|---|---|---|---|---|---|
| Acute Viral Hepatitis — no.(%) | 0 (0) | 8 (1.38) | 3 (1.62) | 1 (1.01) | 4 (1.35) | 0.91 |
| Hepatocellular Carcinoma — no.(%) | 0 (0) | 18 (3.1) | 8 (4.32) | 4 (4.04) | 6 (2.03) | 0.31 |
| Number of clinical events — number | 0 (0) | 2.09 ± 0.97 | 2.7 ± 1 | 2.27 ± 0.81 | 1.66 ± 0.75 | <0.01 |
| Number of precipitating events — number | 0 (0) | 0.42 ± 0.57 | 0.76 ± 0.63 | 0.21 ± 0.41 | 0.28 ± 0.47 | <0.01 |
| Liver Dysfunction— no.(%) * | 0 (0) | 72 (12.41) | 56 (30.27) | 5 (5.05) | 11 (3.72) | <0.01 |
| Renal Dysfunction — no.(%) * | 0 (0) | 61 (10.52) | 26 (14.05) | 12 (12.12) | 23 (7.77) | 0.08 |
| Brain Dysfunction — no.(%) * | 0 (0) | 174 (30) | 81 (43.78) | 93 (93.94) | 0 (0) | <0.01 |
| Coagulation Dysfunction — no.(%) * | 0 (0) | 58 (10) | 45 (24.32) | 6 (6.06) | 7 (2.36) | <0.01 |
| Heart Dysfunction — no.(%) * | 0 (0) | 52 (8.97) | 24 (12.97) | 5 (5.05) | 23 (7.77) | 0.05 |
| Respiratory Dysfunction — no.(%) * | 0 (0) | 31 (5.34) | 16 (8.65) | 5 (5.05) | 10 (3.38) | 0.04 |
| Liver Failure — no.(%) * | 0 (0) | 9 (1.55) | 8 (4.32) | 0 (0) | 1 (0.34) | <0.01 |
| Brain Liver Failure — no.(%) * | 0 (0) | 15 (2.59) | 9 (4.86) | 6 (6.06) | 0 (0) | <0.01 |
| Coagulation Failure — no.(%) * | 0 (0) | 10 (1.72) | 7 (3.78) | 0 (0) | 3 (1.01) | 0.03 |
| Heart Failure — no.(%) * | 0 (0) | 3 (0.52) | 2 (1.08) | 0 (0) | 1 (0.34) | 0.40 |
| Respiratory Failure — no.(%) * | 0 (0) | 3 (0.52) | 0 (0) | 0 (0) | 3 (1.01) | 0.24 |
| Number of dysfunctions — number | 0 (0) | 0.77 ± 0.89 | 1.34 ± 1 | 1.27 ± 0.59 | 0.25 ± 0.49 | 0.00 |
| Number of failures — number | 0 (0) | 0.07 ± 0.25 | 0.14 ± 0.35 | 0.06 ± 0.24 | 0.03 ± 0.16 | <0.01 |
| **Cirrhosis Severity Scores** | | | | | | |
| Child-Pugh | 12 (2.07) | 8.58 ± 1.97 | 10.18 ± 1.37 | 8.8 ± 1.74 | 7.5 ± 1.62 | <0.01 |
| CLIF-C-AD | 20 (3.45) | 50.65 ± 9.03 | 54.6 ± 8.92 | 51.19 ± 8.25 | 48 ± 8.43 | <0.01 |
| CLIF-OF | 0 (0) | 6.81 ± 0.97 | 7.48 ± 1.07 | 7.27 ± 0.68 | 6.23 ± 0.52 | <0.01 |
| MELD | 1 (0.17) | 15.31 ± 4.86 | 19.48 ± 3.9 | 14.56 ± 4.14 | 12.95 ± 3.8 | <0.01 |
| MELD-Na | 3 (0.52) | 18.08 ± 5.51 | 22.56 ± 4.09 | 17.51 ± 4.95 | 15.47 ± 4.64 | <0.01 |
| **Medical History** | | | | | | |
| Diabetes Mellitus — no.(%) | 0 (0) | 228 (39.31) | 12 (6.49) | 63 (63.64) | 153 (51.69) | <0.01 |
| Arterial hypertension — no.(%) | 0 (0) | 193 (33.28) | 38 (20.54) | 47 (47.47) | 108 (36.49) | <0.01 |
| History of previous decompensations — no.(%) | 12 (2.07) | 494 (85.17) | 161 (87.03) | 90 (90.91) | 243 (82.09) | 0.07 |
| **Lifestyle** | | | | | | |
| Alcohol consumption in the 3 previous months — no.(%) | 0 (0) | 224 (38.62) | 83 (44.86) | 35 (35.35) | 106 (35.81) | 0.11 |
| Active smoker — no.(%) | 0 (0) | 195 (33.62) | 67 (36.22) | 26 (26.26) | 102 (34.46) | 0.22 |
| **Laboratory Variables** | | | | | | |
| Serum Alanine Transaminase — U/L | 20 (3.45) | 43.8 ± 48.17 | 51.25 ± 54.79 | 39.04 ± 42.45 | 40.74 ± 45.06 | 0.03 |
| Serum Aspartate aminotransferase — U/L | 22 (3.79) | 67.5 ± 64.76 | 90.85 ± 93.03 | 53.71 ± 27.65 | 57.52 ± 45.54 | <0.01 |
| Serum Albumin — g/dL | 12 (2.07) | 2.92 ± 0.58 | 2.71 ± 0.52 | 2.94 ± 0.6 | 3.05 ± 0.57 | <0.01 |
| Serum Total Bilirubin — mg/dL | 1 (0.17) | 3.09 ± 3.25 | 5.29 ± 4.19 | 2.37 ± 2.08 | 1.96 ± 1.96 | <0.01 |
| Serum Gamma-glutamyl transferase — U/L | 136 (23.45) | 143.55 ± 175.46 | 155.94 ± 222.34 | 123.37 ± 123.06 | 142.55 ± 155.96 | 0.49 |
| Serum C-Reactive Protein — mg/L | 36 (6.21) | 35.45 ± 38.44 | 41.7 ± 40.04 | 32 ± 42.65 | 32.69 ± 35.49 | 0.02 |

| | | | | | | |
|---|---|---|---|---|---|---|
| Serum Sodium— mEq/L | 2 (0.34) | 135.7 ± 5.45 | 134.03 ± 5.77 | 135.72 ± 6.36 | 136.74 ± 4.62 | <0.01 |
| Serum Potassium — mEq/L | 6 (1.03) | 4.19 ± 0.61 | 4.19 ± 0.68 | 4.2 ± 0.56 | 4.18 ± 0.57 | 0.74 |
| Hemoglobin — g/dL | 3 (0.52) | 9.91 ± 2.19 | 10.09 ± 2.1 | 10.2 ± 2.08 | 9.7 ± 2.27 | 0.04 |
| Hematocrit — % | 4 (0.69) | 29.56 ± 6.14 | 29.74 ± 6.1 | 30.28 ± 5.91 | 29.2 ± 6.23 | 0.29 |
| Creatinine — mg/dL | 0 (0) | 0.98 ± 0.36 | 1.03 ± 0.39 | 0.97 ± 0.35 | 0.94 ± 0.33 | 0.01 |
| White blood cell count — x10^9/L | 18 (3.1) | 6.08 ± 3.83 | 7.25 ± 4.64 | 5.39 ± 2.67 | 5.57 ± 3.44 | <0.01 |
| Lymphocytes — x10^9/L | 129 (22.24) | 0.99 ± 0.73 | 1.09 ± 0.89 | 0.92 ± 0.48 | 0.96 ± 0.69 | 0.07 |
| Monocytes — x10^9/L | 165 (28.45) | 0.55 ± 0.42 | 0.62 ± 0.4 | 0.5 ± 0.24 | 0.52 ± 0.48 | 0.01 |
| Neutrophils — x10^9/L | 126 (21.72) | 4.28 ± 3.21 | 5.29 ± 4.04 | 3.74 ± 2.26 | 3.83 ± 2.73 | <0.01 |
| INR (International Normalized Ratio) | 0 (0) | 1.56 ± 0.39 | 1.81 ± 0.45 | 1.53 ± 0.31 | 1.42 ± 0.29 | <0.01 |
| Platelet — x10^3/µL | 5 (0.86) | 104.22 ± 66.73 | 99.62 ± 62.91 | 89.15 ± 45.35 | 112.14 ± 73.73 | 0.03 |
| SpO2 — (%) | 15 (2.59) | 96.24 ± 2.32 | 96.3 ± 2.23 | 96.2 ± 2.45 | 96.22 ± 2.34 | 0.73 |
| SpO2/FiO2 Ratio | 15 (2.59) | 449.3 ± 39.73 | 447.5 ± 42.24 | 447.46 ± 44.5 | 451.03 ± 36.34 | 0.32 |
| **Outcomes** | | | | | | |
| 28-day mortality — no.(%) | 0 (0) | 59 (10.2) | 26 (4.48%) | 13 (2.24) | 20 (3.45) | 0.02 |
| 28-day ACLF development — no.(%) | 0 (0) | 65 (11.2) | 29 (5%) | 16 (2.76) | 20 (3.45) | <0.01 |
| 28-day transplant — no.(%) | 0 (0) | 18 (3.1) | 12 (2.07%) | 3 (0.52) | 3 (0.52) | <0.01 |

*ACLF=Acute-on-chronic liver failure, FiO2= Fraction of inspired oxygen NASH= Non-alcoholic steatohepatitis, SpO2= Saturation of peripheral Oxygen..*

** as defined in Jalan, R., et al. (2014). Development and validation of a prognostic score to predict mortality in patients with acute-on-chronic liver failure. Journal of Hepatology, 61(5), 1038–1047. https://doi.org/10.1016/j.jhep.2014.06.012*

**Table S9**. **Performance measures of kNN classifier.** Summary of the evaluated metrics used to set the choice of the *k* in the kNN model for label transfer.

| k | Accuracy | AUC | FP | FN | ER |
|---|---|---|---|---|---|
| 25 | 0.8304348 | 0.6782492 | 3 | 4 | 0.0304348 |
| 26 | 0.8304348 | 0.6819865 | 3. | 4 | 0.0304348 |
| 27 | 0.8391304 | 0.6814254 | 4 | 4 | 0.0347826 |
| 28 | 0.8304348 | 0.6853535 | 5 | 2 | 0.0304348 |
| 29 | 0.826087 | 0.6886756 | 5 | 3 | 0.0347826 |
| 30 | 0.826087 | 0.6884736 | 4 | 3 | 0.0304348 |

*AUC=Area Under the Curve, FP=False Positives, FN=False Negatives, ER=Error Rate*

***Table S10. Summary of the most predictive variables in the identified clusters in ACLARA cohort at hospital admission.*** The p-values are derived from the ANOVA analysis for continuous variables or from $\chi2$ analysis for binary and categorical variables. Values are the mean ± SD unless indicated otherwise.

| Clinical Feature | Cluster 1 (n=185) | Cluster 2 (n=99) | Cluster 3 (n=296) | P-value |
|---|---|---|---|---|
| Diabetes Mellitus — no.(%) | 12 (6.49) | 63 (63.64) | 153 (51.69) | 2.50E-28 |
| Hepatic Encephalophaty — no.(%) | 90 (48.65) | 99 (100) | 0 (0) | 5.48E-81 |
| Clinical Events — number | 2.7 ± 1 | 2.27 ± 0.81 | 1.66 ± 0.75 | 1.98E-27 |
| Organ Dysfunction — number | 1.34 ± 1 | 1.27 ± 0.59 | 0.25 ± 0.49 | 1.35E-56 |
| Precipitating Events — number | 0.76 ± 0.63 | 0.21 ± 0.41 | 0.28 ± 0.47 | 2.42E-20 |
| Serum Albumine — g/dL | 2.71 ± 0.52 | 2.94 ± 0.6 | 3.05 ± 0.57 | 1.80E-10 |
| Serum Aspartate Aminotransferase — units/L | 90.85 ± 93.03 | 53.71 ± 27.65 | 57.52 ± 45.54 | 1.20E-07 |
| Serum CRP — mg/L | 41.7 ± 40.04 | 32 ± 42.65 | 32.69 ± 35.49 | 1.66E-02 |
| Serum Bilirubin —mg/dL | 5.29 ± 4.19 | 2.37 ± 2.08 | 1.96 ± 1.96 | 3.03E-29 |
| INR | 1.81 ± 0.45 | 1.53 ± 0.31 | 1.42 ± 0.29 | 7.59E-28 |
| Serum Sodium — mEq/L | 134.03 ± 5.77 | 135.72 ± 6.36 | 136.74 ± 4.62 | 9.14E-08 |
| White Blood Cells — $x10^3/mm^3$ | 7.25 ± 4.64 | 5.39 ± 2.67 | 5.57 ± 3.44 | 7.04E-06 |

** CRP=C-Reactive Protein, INR= International Normalized Ratio.*

¶ *Counting the hospital admission visit.*

**Table S11. Comparison of the cumulative incidence models at different time points.** Summary of the evaluated goodness-of-fit metrics to compare the cumulative incidence models based on the stratification at hospital admission *vs* at the last reported visit before any event occurrence in the PREDICT cohort.

| | | N=688 | |
|---|---|---|---|
| | | Hospital Admission | "Last visit" |
| ACLF | Log-rank Test | 3e-04 | 1e-11 |
| | Likelihood ratio | 15.69 | 38.16 |
| | Concordance | 0.613 | 0.665 |
| | BIC | 1021.72 | 994.87 |
| | AIC | 1016.96 | 992.48 |
| Death | Log-rank Test | 2e-04 | 1e-09 |
| | Likelihood ratio | 18.45 | 31.5 |
| | Concordance | 0.621 | 0.646 |
| | BIC | 1060.07 | 1042.59 |
| | AIC | 1055.23 | 1040.18 |

*BIC= Bayesian Information Criterion, AIC=Akaike Information Criterion*
